# BMI Variability and Cardiovascular Outcomes Within Clinical Trial and Real-World Environments in Type 2 Diabetes: An IMI2 SOPHIA study

**DOI:** 10.1101/2024.03.15.24303590

**Authors:** Robert J Massey, Yu Chen, Marina Panova-Noeva, Michaela Mattheus, Moneeza K Siddiqui, Nanette C Schloot, Antonio Ceriello, Ewan R Pearson, Adem Y Dawed

## Abstract

**Aims:** BMI variability has been associated with increased cardiovascular disease risk in individuals with type 2 diabetes, however comparison between clinical studies and real-world observational evidence has been lacking. Furthermore, it is not known whether BMI variability has an effect independent of HbA1c variability.

**Methods and Results:** We investigated the association between BMI variability and 3P-MACE risk in the Harmony Outcomes trial (n = 9198), and further analysed placebo arms of REWIND (n = 4440) and EMPA-REG OUTCOME (n = 2333) trials, followed by real-world data from the Tayside Bioresource (n = 6980) using Cox regression modelling. BMI variability was determined using average successive variability (ASV), with first major adverse cardiovascular event of non-fatal stroke, non-fatal myocardial infarction, and cardiovascular death (3P-MACE) as the primary outcome.

After adjusting for cardiovascular risk factors, a +1 SD increase in BMI variability was associated with increased 3P-MACE risk in Harmony Outcomes (HR 1.12, 95% CI 1.08 – 1.17, P < 0.001). The most variable quartile of participants experienced an 87% higher risk of 3P-MACE (P <0.001) relative to the least variable. Similar associations were found in REWIND and Tayside Bioresource. Further analyses in the EMPA-REG OUTCOME trial did not replicate this association. BMI variability’s impact on 3P-MACE risk was independent of HbA1c variability.

**Conclusion:** In individuals with type 2 diabetes, increased BMI variability was found to be an independent risk factor for 3P-MACE across cardiovascular outcome trials and real-world datasets. Future research should attempt to establish a causal relationship between BMI variability and cardiovascular outcomes.

## 1. Introduction

Repeated cycles of weight loss and weight regain is often referred to as weight “cycling”, “fluctuation”, or “variability”. This variability may be harmful, as a variety of studies have found that variability in clinical biomarkers predict worse outcomes (1–3). Indeed, an increase in weight variability has recently been found to be associated with an increase in the risk of cardiovascular disease and major adverse cardiovascular events (3P-MACE) (4, 5). We recently performed a literature review and meta-analysis which observed an increase in the risk of cardiovascular morbidity and mortality associated with body weight and BMI variability independent of type 2 diabetes status (6). Despite these findings however, the topic of weight variability remains controversial, as not all studies have observed this relationship between weight variability and 3P-MACE (7, 8), whilst others have observed that weight variability predicted lower cardiovascular disease risk (9).

A potential explanation for this observed heterogeneity may be due to differences in the statistical models utilised. These models frequently control for disparate confounding variables or may be inadequately adjusted for important variables. For example, no BMI variability study to date has controlled for the established effect of HbA1c variability on cardiovascular health in individuals with type 2 diabetes (1). Given the intrinsic link between HbA1c and BMI variability, it is possible that the cardiovascular risk signal associated with BMI variability is influenced by HbA1c variability (10). Moreover, while previous analyses have separately examined clinical and observational cohorts, none have explored the impact of BMI variability in both types of cohorts using a standard statistical model. This is important as differences inherent to these cohorts may explain some of the observed variability between studies.

Therefore, the primary aim of this study was to comprehensively investigate the association between BMI variability and the risk of 3-point MACE (non-fatal stroke, non-fatal myocardial infarction, and cardiovascular death) within clinical trial datasets and observational cohorts comprised of individuals with type 2 diabetes using standard statistical models. The secondary aim of this study was to investigate if BMI variability and glycaemic variability are independent risk factors for 3P-MACE. A Cox proportional hazards model was created to assess the association between BMI variability and 3-point MACE using the Harmony Outcomes trial as a discovery dataset; this model was then used to perform separate analyses within the placebo arms of the Researching cardiovascular Events with a Weekly Incretin in Diabetes (REWIND) and Empagliflozin Cardiovascular Outcome Event Trial in Type 2 Diabetes Mellitus Patients–Removing Excess Glucose (EMPA-REG) OUTCOME trials. We then used the same model for a further analysis within the real-world observational data collected from the Genetics of Diabetes Audit and Research Tayside and Scotland (GoDARTS) and the Scotland Health Research Register (SHARE) cohorts, combined into a single cohort referred to as Tayside Bioresource.

## 2. Methods

### 2.1. Study Design and Participants

Trial data sets: Post-hoc analyses of trial data from the Harmony Outcomes consisting of 9,463 participants was used as a discovery cohort. First round replication was performed using the placebo arm of the REWIND study (n = 4440) and final replication attempt was performed using data from the placebo arm of the EMPA-REG OUTCOME trial (n = 2333). All three trials, involving patients with type 2 diabetes, were international multi-centre, randomised, double-blind, placebo-controlled studies that independently investigated the effect of antidiabetic medications on 3P-MACE risk; Harmony Outcomes and REWIND investigated the efficacy of glucagon-like peptide-1 receptor agonists (GLP-1RAs) albiglutide and dulaglutide, respectively, whereas EMPA-REG OUTCOME investigated the efficacy of sodium-glucose cotransporter-2 inhibitor (SGLT2i) empagliflozin. Detailed descriptions of the included clinical cohorts can be found in the supplementary material (Appendix 1).

Real-world data analysis: A retrospective follow-up survival study was performed among participants from a combination of observational cohorts from the GoDARTS and the SHARE studies. Further details on these cohorts can be found in the supplementary material (Appendix 1). Electronic health records (from primary and secondary care), biochemistry, prescribing records, and data from the Scottish death register are pooled from the Scottish Care Information – Diabetes Collaboration, GoDARTS, and SHARE to form the Tayside Bioresource. Data collected between January 1^st^ 1994 to January 1^st^ 2020 were considered for this study. Entry to study for each individual was either the date of 1^st^ recorded BMI measurement or date of type 2 diabetes diagnosis, whichever was latest. Our analysis of Tayside Bioresource consisted of 2 phases: a 5 year exposure phase where we calculated BMI variability and other baseline variables, followed by a 10 year longitudinal phase for the observation of 3P-MACE where the Cox regression analysis was performed. We identified individuals with type 2 diabetes that had at least 3 recorded measurements of BMI during the exposure phase. Participants that experienced a primary outcome event during this exposure phase were removed from the study as we believed this would impact calculated BMI variability. Similarly, patients recorded as pregnant or having undergone a limb amputation during these five years were also excluded. Patients meeting these criteria entered the longitudinal phase of the study where they were then followed either until they experienced a primary outcome event or their final BMI measurement within the 10 years of follow-up; whichever came first. If an individual’s 10-year follow-up period extended beyond January 1st, 2020, they were excluded from the analysis. This meant the latest possible entry to the study was January 1st, 2005; individuals entering the study after this date could not undergo the full 15 years of analysis and were thus removed.

### 2.2. Study Outcomes

The primary outcomes of Harmony Outcomes, REWIND, EMPA-REG OUTCOME, and Tayside Bioresource were similar, each defined as the first occurrence of 3-point MACE, which was a composite of cardiovascular death, non-fatal myocardial infarction, and non-fatal stroke (fatal and non-fatal in Harmony Outcomes). In our study, we defined the primary outcome as the first occurrence of 3-point MACE, as defined in each study by their respective study designs (11–13).

The primary outcome of the analysis of Tayside Bioresource was also defined as 3-point MACE, the first incidence of non-fatal myocardial infarction, non-fatal stroke, and cardiovascular mortality.

### 2.3. Measurement of BMI Variability

For post-hoc analyses, BMI variability was defined as the fluctuation in BMI across visits, measured using average successive variability (ASV). In Harmony Outcomes, REWIND, and EMPA-REG OUTCOME, at least 3 BMI measurements per person were required for ASV calculation. ASV was based on all available data until 3P-MACE occurrence or censoring.

In Tayside Bioresource analysis, BMI variability was defined similarly but adjusted for irregular visit patterns by dividing the absolute difference between BMI measurements by the time interval between them. A minimum 90-day interval between BMI measurements and at least 3 recorded measurements per person were required for inclusion.

### 2.4. Statistical Analyses

The association between BMI variability as calculated by ASV and the risk of 3P-MACE outcomes was analysed by considering body-weight variability as both a continuous and categorical variable. Within the trial cohorts, any individuals that experienced 3P-MACE events before 4 months of follow up were excluded as this was the minimum date at which BMI variability could be calculated.

To analyse the risk of 3P-MACE associated with BMI variability as a continuous variable within the Harmony Outcomes trial, 5 Cox proportional-hazards regression models were created to estimate the hazard ratios (HR) and 95% confidence intervals. Each model investigated the HR of 3P-MACE associated with an increase of ASV BMI by 1 standard deviation (SD). Model 1 was unadjusted; model 2 included trial arm (treatment vs placebo) as a covariate; model 3 added baseline BMI to model 2 as a covariate; model 4 further included baseline age, sex, type 2 diabetes duration at baseline, smoking history, baseline systolic blood pressure, statin use, baseline HbA1c, the number of BMI measures, and baseline cardiovascular risk to model 3; model 5 was then created with ASV of HbA1c (+1 SD) added to model 4. This was primarily to test whether the 3P-MACE risk associated with BMI variability was independent of the 3P-MACE risk associated with HbA1c variability. The analyses of models 4 and 5 were then created within the REWIND, EMPA-REG OUTCOME, and Tayside Bioresource cohort data, with a few adjustments due to data availability: in REWIND, the covariate for history of previous cardiovascular disease was included instead of baseline cardiovascular risk, and type 2 diabetes duration at baseline was not included; for the EMPA-REG OUTCOME trial data the models were constructed with a covariate for lipid controlling drug use at baseline instead of statin use at baseline; the models constructed within the Tayside Bioresource cohort did not include covariates for cardiovascular risk, statin use, or baseline systolic blood pressure, but instead included covariates for lipid-controlling and antihypertensive drug use. Additionally, the age covariate within the models constructed within the Tayside Bioresource cohort was split into quartiles in order to allow the models to meet the proportional hazards assumption.

To analyse the risk of 3P-MACE associated with BMI variability as a categorical variable, participants were divided into quartiles based on calculated ASV BMI values. Cox proportional-hazards regression analyses were performed to estimate the relative risk of 3P-MACE of each quartile of BMI variability compared to the lowest quartile.

Further analyses were performed using continuous BMI variability in order to investigate the effect of baseline BMI on the risk relationship observed between BMI variability and 3P-MACE. We assigned individuals in each trial to one of 3 groups: normal baseline BMI (≤25 kg/m^2^); overweight (>25 to ≤30 kg/m^2^); and obese (>30 kg/m^2^). Cox proportional-hazards regression analyses (fitting model 5 as above) were then performed to estimate the risk of 3P-MACE per +1 SD increase in ASV BMI.

Finally, we performed two meta-analyses of trial results using summary statistics from Harmony Outcomes, REWIND, and EMPA-REG OUTCOME, for continuous and categorical BMI variability. We then performed an additional two meta-analyses for continuous and categorical BMI variability, this time including Tayside Bioresource. For continuous BMI variability, this was done by taking the HR estimates for +1 SD increase in ASV BMI from model 5 of each cohort and pooling them via a fixed effects model. The meta-analysis of categorical BMI variability was conducted by the same methodology, whereby HR estimates for the most fluctuating quartiles from model 4 of each were combined. These meta-analyses were performed using the ‘metafor’ package in R (version 4.1.1).

### 2.5. Data availability statement

The datasets generated during and/or analysed in the current study are available from the corresponding author upon reasonable request.

## 3. Results

### 3.1. Population Characteristics

The study population sizes from the Harmony Outcomes and the placebo arms of REWIND, and EMPA-REG OUTCOME trials were 9198, 4440, and 2171, respectively. The number of individuals from the Tayside Bioresource cohort that met inclusion criteria for the current investigation was 6980. The clinical features of the participants of each of these 4 cohorts are listed in Table S1.

### 3.2. BMI variability and 3P-MACE

In our post-hoc analysis of the discovery cohort from the Harmony Outcomes 6.75% of participants experienced a 3P-MACE event (n = 621). When BMI variability was considered as a continuous covariate in the fully adjusted model (model 4), an increase in ASV BMI of 1 SD was associated with an increase in the risk of a 3P-MACE outcome (HR 1.12, 95% confidence intervals (CI) 1.08 – 1.17, P < 0.001; Figure 1a). This was independent of traditional cardiovascular risk factors. Furthermore, this effect was independent of GLP1RA drug use, where albiglutide treatment was still associated with decreased 3P-MACE risk (HR 0.70, 95% CI 0.60 – 0.82, P <0.001). When BMI variability was treated as a categorical variable, individuals in the top quartile of BMI variability compared to individuals in the lowest quartile of variability had an increased risk of 3P-MACE outcomes (HR 1.87, 95% CI 1.49 – 2.36, P < 0.001; Figure S1a).

**Figure 1(a):**
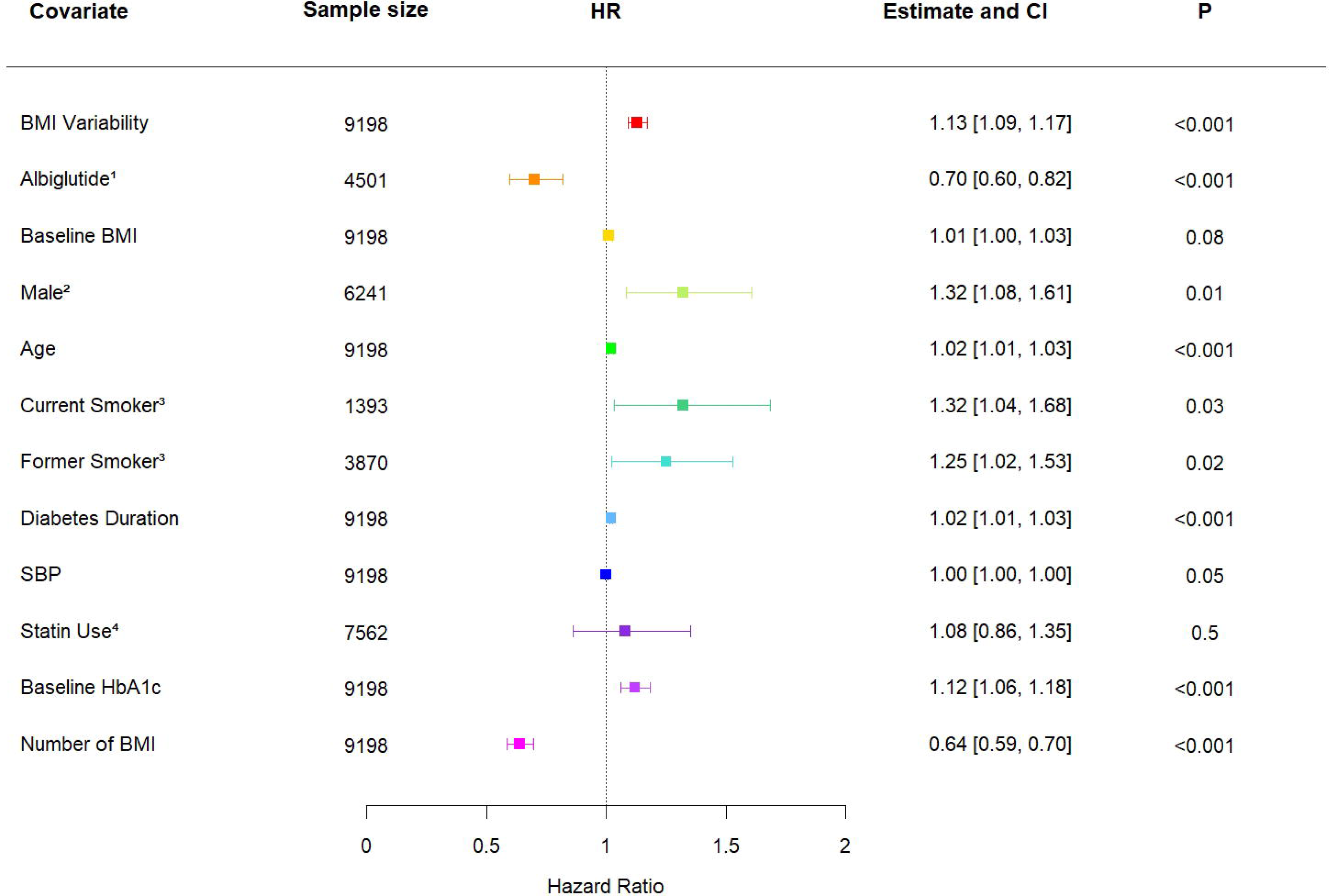
A forest plot summarising the adjusted hazard ratio (HR) and 95% confidence interval (CI) of 3-point major adverse cardiovascular events (3P-MACE) risk associated with a +1 standard deviation (SD) increase in BMI variability within the Harmony Outcomes trial cohort (n = 9198) after adjustment for treatment, baseline BMI, sex, age, smoking, type 2 diabetes duration, systolic blood pressure, statin use, baseline HbA1c, and number of BMI measurement. Risk estimates (HR and 95% confidence interval (CI)) presented based on Cox regression model versus reference for categorical variables and for a one-unit increase for continuous variables. ^1^ reference Placebo, n = 4473; ^2^ reference Female, n = 2733; ^3^ reference Never Smoked, n = 3710; ^4^ reference No Statin Use, n = 1411.

When we performed this analysis within the placebo arm of the REWIND trial, we found similar results. In this cohort, 11.58% experienced a 3P-MACE outcome (n = 514). When BMI variability was included into model 4 as a continuous covariate, we found that an increase in ASV BMI of 1 SD was associated with an increase in 3P-MACE risk (HR 1.09, 95% CI 1.02 – 1.16, P = 0.016; Figure 1b). This increase was again independent of the 3P-MACE risk associated with classic cardiovascular risk factors. Treating BMI variability categorically, there was a significantly increased risk of 3P-MACE for individuals in the top quartile of BMI variability compared to the lowest quartile (HR 1.95, 95% CI 1.50 – 2.54, P <0.001; Figure S1b).

**Figure 1(b):**
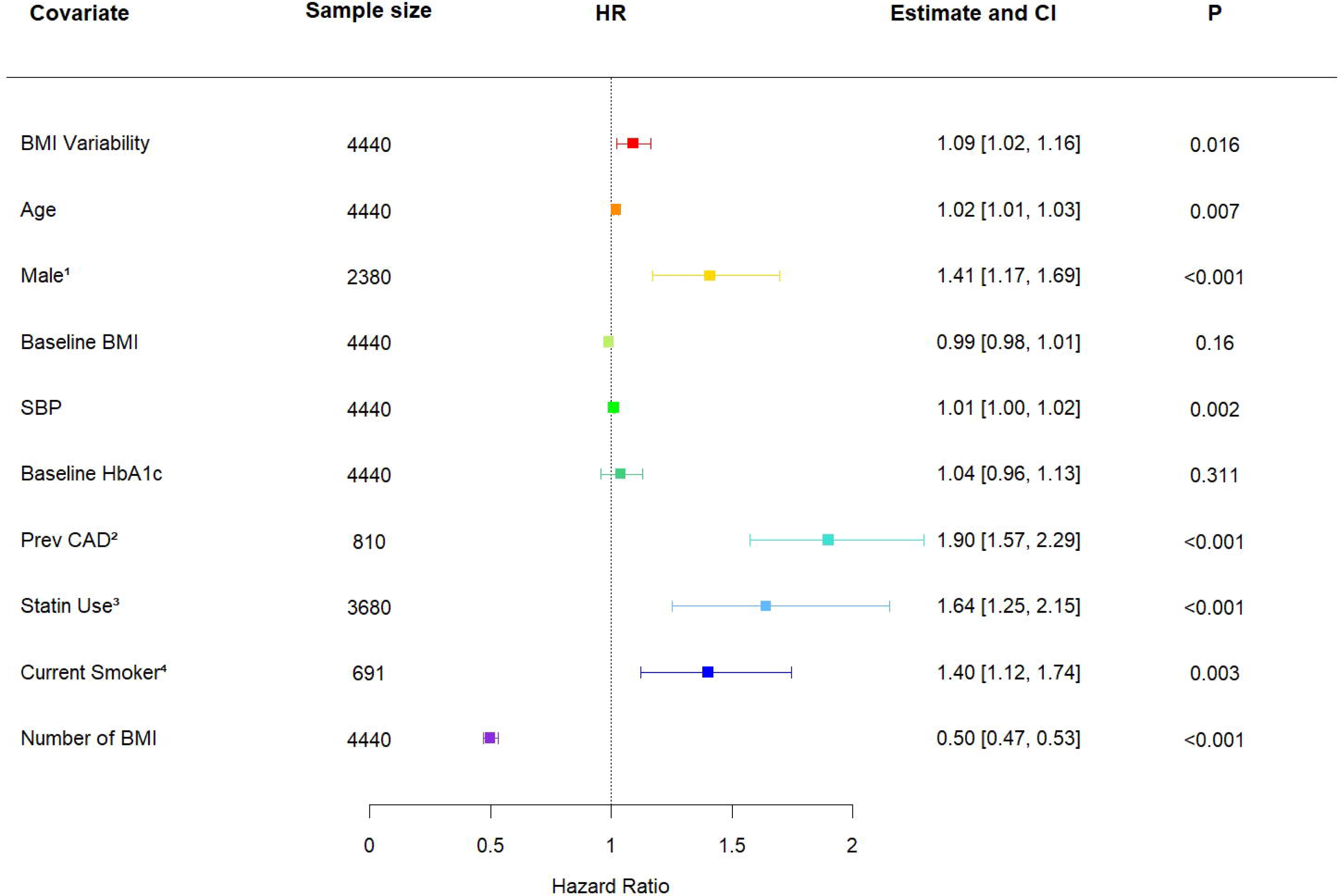
A forest plot summarising the adjusted hazard ratio (HR) and 95% confidence interval (CI) of 3-point major adverse cardiovascular events (3P-MACE) risk associated with a +1 standard deviation (SD) increase in BMI variability within the REWIND trial placebo-arm cohort (n = 4440) after adjustment for age, sex, baseline BMI, systolic blood pressure, baseline HbA1c, history of coronary artery disease (CAD), statin use, smoking, and number of BMI measurement. Risk estimates (HR and 95% confidence interval (CI)) presented based on Cox regression model versus reference for categorical variables and for a one-unit increase for continuous variables. ^1^ reference Female, n = 2060; ^2^ reference No CAD, n = 3630; ^3^ reference No Statin Use, n = 760; ^4^ reference No Smoking, n = 3749.

We then performed our analysis within the placebo arm of the EMPA-REG OUTCOME trial. In this cohort, 9.40% experienced a 3P-MACE outcome (n = 204). When BMI variability was included into model 4 as a continuous variable, an increase in ASV BMI of +1 SD was shown to be associated with a decrease in 3P-MACE risk (HR 0.80, 95% CI 0.65 – 0.97, P = 0.0251; Figure 1c). This association was independent of the classic 3P-MACE risk factors included into the model. Similarly, when BMI variability was considered as a categorical variable, the individuals in the most fluctuating quartile experienced a significantly decreased risk of 3P-MACE (HR 0.61, 95% CI 0.42 – 0.90, P = 0.0115; Table S2).

**Figure 1(c):**
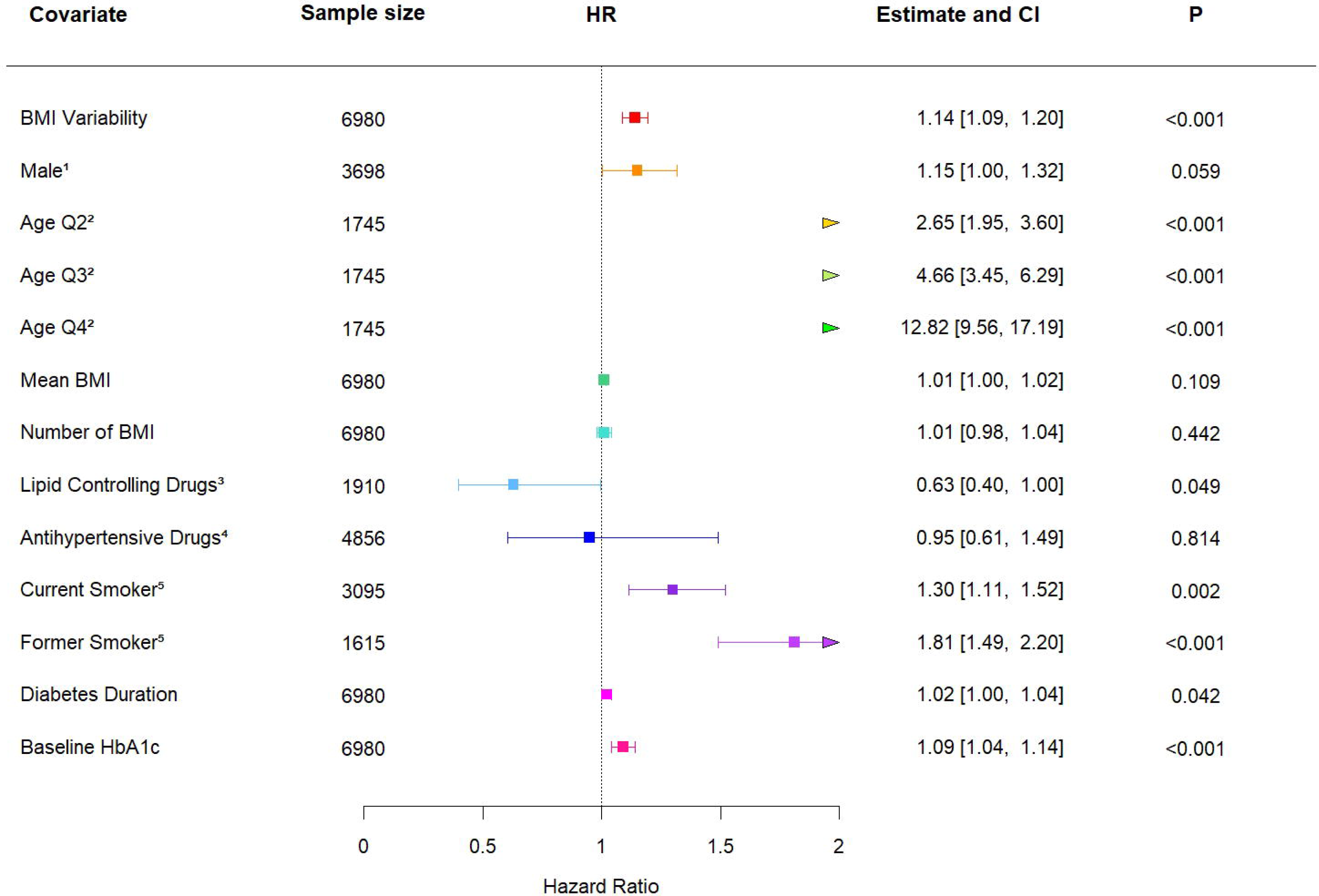
A forest plot summarising the adjusted hazard ratio (HR) and 95% confidence interval (CI) of 3-point major adverse cardiovascular events (3P-MACE) risk associated with a +1 standard deviation (SD) increase in BMI variability within the EMPA-REG OUTCOME trial placebo-arm cohort (n = 2171) after adjustment for baseline BMI, systolic blood pressure, diastolic blood pressure, sex, age, number of BMI measurements, lipid controlling drug use, smoking status, type 2 diabetes duration, and baseline HbA1c. Risk estimates (HR and 95% confidence interval (CI)) presented based on Cox regression model versus reference for categorical variables and for a one-unit increase for continuous variables. ^1^ reference Male, n = 2060; ^2^ reference No Lipid Controlling Drugs, n = 3630; ^3^ reference No Smoking, n = 760; ^4^ reference Diabetes Duration x<1, n = 3749. Diabetes duration is split in to categories: less than 1 year (x<1); between 1 and 5 years (1<x≤5); between 5 and 10 years (5<x≤10); over 10 years (x>10).

Finally, we performed our analysis with the Tayside Bioresource cohort. Among the 6980 individuals in this cohort, 11.60% experienced a 3P-MACE outcome (n = 812). When BMI variability was included into model 4 as a continuous variable, an increase in ASV BMI of +1 SD was associated with an increase in the risk of a 3P-MACE event (HR 1.14, 95% CI 1.09 – 1.20, P <0.001; Figure 1d) after controlling for classic cardiovascular risk factors. When BMI variability was treated as a categorical variable, the risk of 3P-MACE for individuals in the most fluctuating quartile was significantly increased (HR 1.47, 95% CI 1.18 – 1.80, P < 0.001; Figure S1c) relative to the least fluctuating quartile.

**Figure 1(d):**
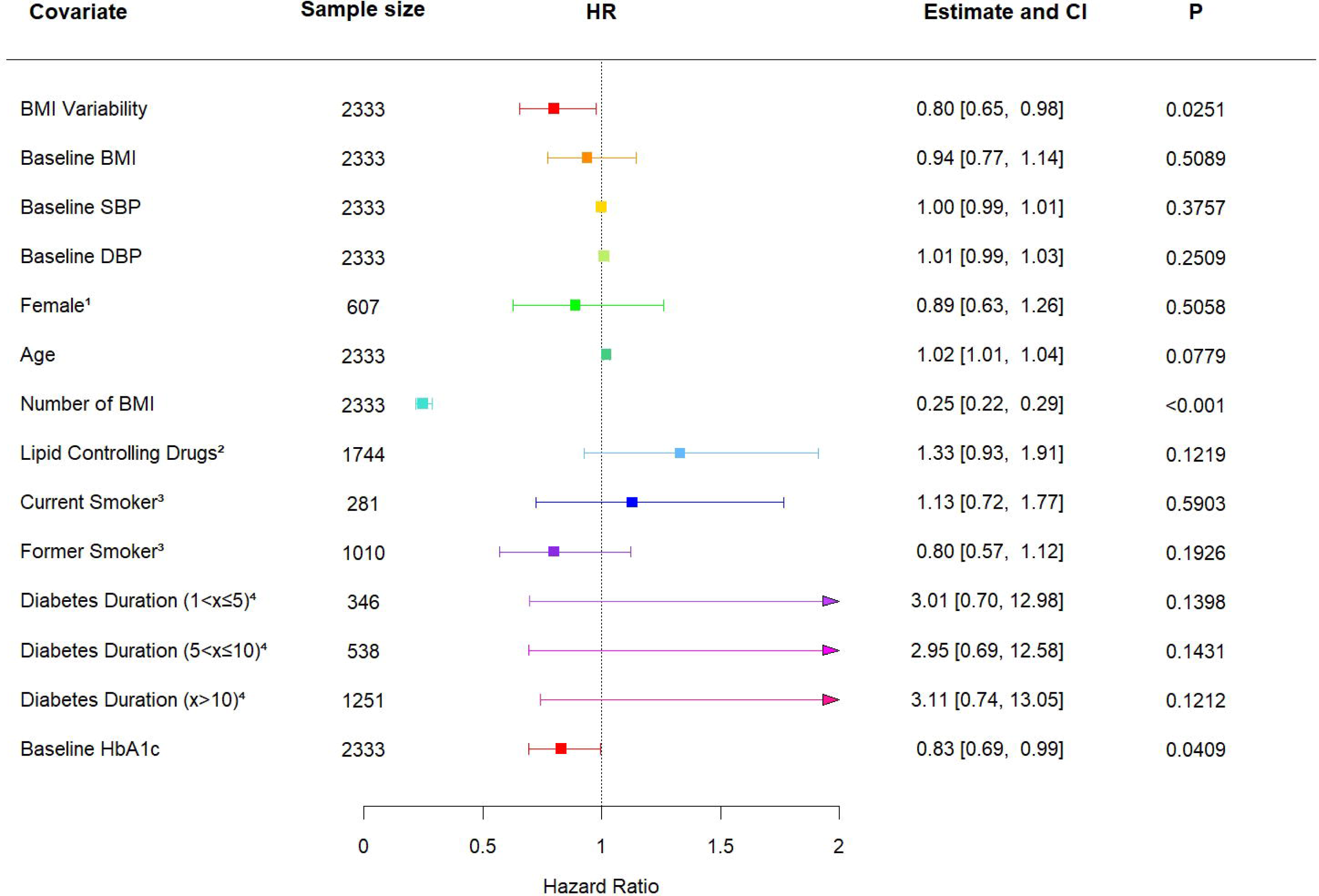
A forest plot summarising the adjusted hazard ratio (HR) and 95% confidence interval (CI) of 3-point major adverse cardiovascular events (3P-MACE) risk associated with a +1 standard deviation (SD) increase in BMI variability within the Tayside Bioresource cohort (n = 6980) after adjustment for sex, age, mean BMI, number of BMI measurements used, lipid-controlling drug use, antihypertensive drug use, smoking status, type 2 diabetes duration, and baseline HbA1c. The covariate “age” has been split into quartiles for this model in order to allow the model to meet proportional hazards assumptions. Risk estimates (HR and 95% confidence interval (CI)) presented based on Cox regression model versus reference for categorical variables and for a one-unit increase for continuous variables. ^1^ reference Female, n = 3282; ^2^ reference Age Quartile 1, n = 1745; ^3^ reference No Lipid Controlling Drug Use, n = 5070; ^4^ reference antihypertensive drug use, n = 2124; ^5^ reference Never Smoked, n = 2270.

### 3.3. Baseline BMI, BMI variability, and 3P-MACE

The majority of participants of all cohorts were either overweight or obese. In the Harmony Outcomes trial, 90.01% (n = 8287) of individuals had a BMI >25 kg/m^2^; within REWIND, 93.53% (n = 4153); within EMPA-REG OUTCOME, 87.52% (n = 1900); and within the Tayside Bioresource cohort, 88.17% participants (n = 5867) were either overweight or obese.

When stratifying participants in Harmony Outcomes, REWIND, EMPA-REG OUTCOME, and Tayside Bioresource into normal weight, overweight, or obese categories, we found that baseline BMI did not appear to modify the association between BMI variability and 3P-MACE risk. Detailed results of the statistical analyses can be found in Appendix 2.

### 3.4. HbA1c variability, BMI variability, and 3P-MACE

In the Harmony Outcomes cohort, a +1 SD increase in ASV HbA1c was associated with an increase in 3P-MACE risk (HR 1.14, 95% CI 1.03 – 1.27, P = 0.014; Figure S2(a)) and this increase in risk was independent from the increase in risk associated with a +1 SD increased in ASV BMI (HR 1.20, 95% CI 1.12 – 1.29, P < 0.001; Figure S2(a)). When we analysed the REWIND cohort we observed that a +1 SD increase in ASV HbA1c was not associated with an increase in 3P-MACE risk (HR 0.99, 95% CI 0.86 – 1.15, P = 0.924; Figure S2(b)), however the inclusion of ASV HbA1c as a covariate did not attenuate the observed 3P-MACE risk associated with a +1 SD increase in ASV BMI (HR 1.09, 95% 1.02 – 1.16, P = 0.016; Figure S2(b)). When this analysis was performed in the EMPA-REG OUTCOME cohort, a +1 SD increase in HbA1c was not associated with an increase in 3P-MACE risk (HR 1.04, 0.93 – 1.17, P = 0.4764; Table S3), and this was independent of an association between a +1 SD increase in ASV BMI and significantly decreased 3P-MACE risk (HR 0.80, 95% CI 0.66 – 0.97, P = 0.0261;Table S3). Within the real-world data of the Tayside Bioresource cohort however, a +1 SD in the variability of HbA1c was again associated with significantly increased risk (HR 1.05, 95% CI 1.01 – 1.10, P = 0.014; Figure S2(c)), independent of the risk associated with a +1 SD increase in BMI variability (HR 1.14, 95% CI 1.09 – 1.20, P <0.001; Figure S2(c)).

### 3.5. Meta-analysis of BMI variability and 3P-MACE outcomes

To investigate the overall effect of BMI variability on 3P-MACE risk we performed 2 separate meta-analyses using effect estimates from model 5 in each trial. The first meta-analysis investigated 3P-MACE risk associated with BMI variability as a continuous variable. The second meta-analysis investigated 3P-MACE risk associated with BMI variability treated as a categorical variable. These meta-analyses involved a total of 15,809 participants of whom 1,339 (8.47%) experienced a 3P-MACE outcome. We then performed 2 more meta-analyses identical to the meta-analyses described above with the addition of the summary effect estimate from model 5 of Tayside Bioresource. These latter two meta-analyses involved a total of 22,789 participants of whom 2,151 (9.45%) experienced a 3P-MACE outcome.

The meta-analysis of BMI variability as a continuous variable within Harmony Outcomes, REWIND, and EMPA-REG OUTCOME found that an increase in ASV BMI by +1 SD was significantly associated with an increase in the summary risk estimate of 3P-MACE (HR 1.12, 95% CI 1.07 – 1.17, P <0.001; Figure S8), with significant heterogeneity observed (I^2^ = 87.50%; P for heterogeneity <0.001). The meta-analysis of BMI variability as a categorical measure found that individuals in the top quartile of BMI variability compared to those in the least variable quartile had a significantly increased summary risk estimate of 3P-MACE (HR 1.56, 95% CI 1.33 – 1.83, P <0.001; Figure S7), with this meta-analysis again showing significant heterogeneity (I^2^ = 92.50%; P for heterogeneity <0.001).

The inclusion of Tayside Bioresource into the meta-analysis caused little adjustment to these results: an increase in ASV BMI by +1 SD was significantly associated with an increase in the summary risk estimate of 3P-MACE (HR 1.13, 95% CI 1.09 – 1.17, P <0.001; Figure S9), with significant heterogeneity observed (I^2^ = 81.80%; P for heterogeneity <0.001). The meta-analysis of BMI variability as a categorical measure found that individuals in the top quartile of BMI variability compared to those in the least variable quartile had a significantly increased summary risk estimate of 3P-MACE (HR 1.52, 95% CI 1.34 – 1.73, P <0.001; Figure S10), with this meta-analysis again showing significant heterogeneity (I^2^ = 88.90%; P for heterogeneity <0.001).

## 4. Discussion

In our post-hoc survival analyses of the participants from three trials and a real-world observational cohort, we found that BMI variability was significantly associated with an increase in the risk of 3P-MACE. These observed associations were independent of variability in HbA1c and other classic cardiovascular risk factors.

Our results add to the growing body of evidence that treatment plans for individuals with type 2 diabetes may also need to consider the risk to cardiovascular health contributed by weight variability. Several studies have previously investigated the effect of weight variability on cardiovascular health within individuals with type 2 diabetes in a variety of demographics (4, 14-19). The majority of these studies also observed the association between weight variability and cardiovascular disease, however, some studies found contradictory results. We have previously proposed that this contradiction may be attributed to discrepancies in cohorts and methodology. Heterogeneity in cohort demographic, type of cohort analysed (i.e. clinical trial versus observational), data collection, definition of cardiovascular events, and the definition of weight variability (e.g. coefficient of variation, SD, ASV, etc.) may lead to heterogeneity in observation (6). One novel finding of our study is that this heterogeneity does not appear to be caused by any inherent difference between trial and observational cohorts, as the results seen in our analyses of Harmony Outcomes, REWIND, and Tayside Bioresource were similar. Furthermore, the similarity of these results provides reinforcement to the idea that heterogeneity in the literature may be in large part due to differences in how BMI variability is defined as well as the covariates controlled for by the statistical models employed. However, despite almost identical methodology used for our analyses, we did not see a replication of our results within the EMPA-REG OUTCOME cohort. The reason for this discrepancy is unclear; there are few differences between the populations of each cohort, and each analysis was adjusted for the same covariates that are typical predictors of cardiovascular disease. Our results suggest that inter-study heterogeneity in the association between BMI variability and 3P-MACE risk is not sufficiently explained by differences between cohorts and methodology, and that further research into this phenomenon may reveal some other explanatory variable.

HbA1c variability has been previously observed to increase cardiovascular disease risk (1, 20, 21). It is known that BMI and type 2 diabetes, as well as glycaemic control, are strongly correlated (22–24). It therefore stands to reason that the increased cardiovascular risk associated with BMI variability may instead be generated by HbA1c variability. However, to our knowledge, no one has yet investigated the contribution of HbA1c variability to the increased risk of cardiovascular events associated with weight variability. Whilst we did also find that HbA1c variability is associated with increased cardiovascular risk among the majority of our cohorts, inclusion of HbA1c variability into our fully adjusted models did not attenuate the estimated risk contributed by BMI variability. This would suggest that BMI variability is a risk factor for cardiovascular events independent of HbA1c variability, as well as other classic cardiovascular risk factors. Indeed, our adjusted models suggest that an increase in BMI variability contributes similar cardiovascular risk estimate to that associated with a history of smoking, of the need for lipid-controlling drugs, being male, and having elevated HbA1c. This highlights the clinical relevance of BMI variability, as well as the necessity of further research into how BMI variability develops and how it can be effectively managed or controlled. Treatment of type 2 diabetes in the future may need to take BMI and HbA1c variabilities into consideration.

A further question our research addresses is whether baseline BMI modulates the cardiovascular risk associated with weight variability. Previous studies performed by Bangalore et al. have observed that obese individuals experience a greater cardiovascular risk associated with weight fluctuations when compared to individuals of a normal weight (4, 15). Our analysis of the Harmony Outcomes and REWIND cohorts found no such association: weight variability was associated with a similar 3P-MACE risk estimate in obese, overweight, and normal weight populations, with no real trend in risk estimates observed. This was also true within our analysis of the Tayside Bioresource cohort. Furthermore, our analysis of the EMPA-REG OUTCOME cohort found no difference in the association of weight variability and MACE across normal weight, overweight, and obese individuals. These findings are consistent with the results of our previously published meta-analysis which also observed that in the literature baseline BMI did not modify the association between BMI variability and cardiovascular events (6).

There are a few limitations to our current study. The key limitation of this and other studies is that we have only investigated an association between weight variability and cardiovascular events; from this data we cannot prove causation. While our current study has gone further to show that the increased cardiovascular risk associated with weight variability is independent of other cardiovascular risk factors, it is still possible that an unmeasured variable could cause the increased risk. Another limitation persistent in studies investigating the relationship between weight variability and cardiovascular events, and a limitation to the present study, is the impact of intentionality of weight loss on cardiovascular disease. While it could be argued that the majority of individuals with type 2 diabetes will be intentionally trying to lose weight, and thus intentionality may have little effect on the cardiovascular risk observed, our current research cannot state this conclusively. Further, in the assessment of BMI variability we do not differentiate between weight loss and weight gain, which could both contribute to an increase in variability, and both may have different effects on the risk of cardiovascular events. Similarly, we cannot conclusively state that medication has no effect of body weight variability and cardiovascular risk. It is possible for instance that the sickest patients changed their glycaemic and cardiovascular care more frequently, resulting in greater body weight variability with or without change in cardiovascular risk.

## 5. Conclusions

Our analyses found that variability in BMI is associated with increase in the risk of 3P-MACE outcomes to a similar extent in both clinical and observational cohorts. This effect appears to be independent of HbA1c variability and other traditional cardiovascular risk factors. Further research into establishing a causative relationship between weight variability and cardiovascular events is recommended.

## Data Availability

All data produced in the present study are available upon reasonable request to the authors.

## 6. Acknowledgments

Personal Thanks: The authors would like to personally thank all members of the SOPHIA consortium for their input and helpful discussions during the development of this study. We would also like to thank Enrique Soto, University of Dundee, UK, for their help in the development and QC of the statistical models used for these analyses. Dr. Soto received no financial support for his participation.

## Funding and assistance

This manuscript is part of a Stratification of Obesity Phenotypes to Optimize Future Obesity Therapy (SOPHIA) project (www.imisophia.eu). SOPHIA has received funding from the Innovative Medicines Initiative 2 Joint Undertaking under grant agreement No. 875534. This Joint Undertaking support from the European Union’s Horizon 2020 research and innovation program and EFPIA and T1D Exchange, JDRF, and Obesity Action Coalition.

## Author contributions and guarantor statement

AD conceived the original hypothesis and planned the framework of the presented analysis. AD, RM, YC, and MM performed the computational analyses of the Harmony Outcome, Tayside Bioresource, REWIND, and EMPA-REG OUTCOME cohorts, respectively. The results presented were discussed with MKS, EP, NCS, AC, and MPN, and all authors contributed to the final manuscript. AD assumes responsibility for the integrity and accuracy of the analyses within this article. The communication reflects the author’s view and neither the IMI nor the European Union, EFPIA, or any Associated Partners are responsible for any use that may be made of the information contained therein. All authors have read and approved the final manuscript.

## References

1. Ceriello A, Lucisano G, Prattichizzo F, La Grotta R, Franzén S, Svensson A-M, et al. HbA1c variability predicts cardiovascular complications in type 2 diabetes regardless of being at glycemic target. Cardiovascular Diabetology. 2022;21(1):13.

2. Cohen AA, Leung DL, Legault V, Gravel D, Blanchet FG, Côté AM, et al. Synchrony of biomarker variability indicates a critical transition: Application to mortality prediction in hemodialysis. iScience. 2022;25(6):104385.

3. Ceriello A, Prattichizzo F. Variability of risk factors and diabetes complications. Cardiovasc Diabetol. 2021;20(1):101.

4. Bangalore S, Fayyad R, DeMicco DA, Colhoun HM, Waters DD. Body Weight Variability and Cardiovascular Outcomes in Patients With Type 2 Diabetes Mellitus. Circ Cardiovasc Qual Outcomes. 2018;11(11):e004724.

5. Nam GE, Kim W, Han K, Lee C-W, Kwon Y, Han B, et al. Body Weight Variability and the Risk of Cardiovascular Outcomes and Mortality in Patients With Type 2 Diabetes: A Nationwide Cohort Study. Diabetes care. 2020;43(9):2234–41.

6. Massey RJ, Siddiqui MK, Pearson ER, Dawed AY. Weight variability and cardiovascular outcomes: a systematic review and meta-analysis. Cardiovasc Diabetol. 2023;22(1):5.

7. Cho IJ, Chang HJ, Sung JM, Yun YM, Kim HC, Chung N. Associations of changes in body mass index with all-cause and cardiovascular mortality in healthy middle-aged adults. PLoS One. 2017;12(12):e0189180.

8. Sponholtz TR, van den Heuvel ER, Xanthakis V, Vasan RS. Association of Variability in Body Mass Index and Metabolic Health With Cardiometabolic Disease Risk. J Am Heart Assoc. 2019;8(7):e010793.

9. Jeong S, Choi S, Chang J, Kim K, Kim SM, Hwang SY, et al. Association of weight fluctuation with cardiovascular disease risk among initially obese adults. Scientific Reports. 2021;11(1):10152.

10. Boye KS, Lage MJ, Shinde S, Thieu V, Bae JP. Trends in HbA1c and Body Mass Index Among Individuals with Type 2 Diabetes: Evidence from a US Database 2012-2019. Diabetes Ther. 2021;12(7):2077–87.

11. Gerstein HC, Colhoun HM, Dagenais GR, Diaz R, Lakshmanan M, Pais P, et al. Design and baseline characteristics of participants in the Researching cardiovascular Events with a Weekly INcretin in Diabetes (REWIND) trial on the cardiovascular effects of dulaglutide. Diabetes Obes Metab. 2018;20(1):42–9.

12. Green JB, Hernandez AF, D’Agostino RB, Granger CB, Janmohamed S, Jones NP, et al. Harmony Outcomes: A randomized, double-blind, placebo-controlled trial of the effect of albiglutide on major cardiovascular events in patients with type 2 diabetes mellitus-Rationale, design, and baseline characteristics. Am Heart J. 2018;203:30–8.

13. Zinman B, Inzucchi SE, Lachin JM, Wanner C, Ferrari R, Fitchett D, et al. Rationale, design, and baseline characteristics of a randomized, placebo-controlled cardiovascular outcome trial of empagliflozin (EMPA-REG OUTCOME™). Cardiovascular Diabetology. 2014;13(1):102.

14. Aucott LS, Philip S, Avenell A, Afolabi E, Sattar N, Wild S. Patterns of weight change after the diagnosis of type 2 diabetes in Scotland and their relationship with glycaemic control, mortality and cardiovascular outcomes: a retrospective cohort study. BMJ Open. 2016;6(7):e010836.

15. Bangalore S, Fayyad R, Laskey R, DeMicco DA, Messerli FH, Waters DD. Body-Weight Fluctuations and Outcomes in Coronary Disease. New England Journal of Medicine. 2017;376(14):1332–40.

16. Ceriello A, Lucisano G, Prattichizzo F, Eliasson B, Franzén S, Svensson A-M, et al. Variability in body weight and the risk of cardiovascular complications in type 2 diabetes: results from the Swedish National Diabetes Register. Cardiovascular Diabetology. 2021;20(1):173.

17. Nam GE, Kim W, Han K, Lee CW, Kwon Y, Han B, et al. Body Weight Variability and the Risk of Cardiovascular Outcomes and Mortality in Patients With Type 2 Diabetes: A Nationwide Cohort Study. Diabetes Care. 2020;43(9):2234–41.

18. Lee HJ, Choi EK, Han KD, Kim DH, Lee E, Lee SR, et al. High variability in bodyweight is associated with an increased risk of atrial fibrillation in patients with type 2 diabetes mellitus: a nationwide cohort study. Cardiovasc Diabetol. 2020;19(1):78.

19. Yeboah P, Hsu FC, Bertoni AG, Yeboah J. Body Mass Index, Change in Weight, Body Weight Variability and Outcomes in Type 2 Diabetes Mellitus (from the ACCORD Trial). Am J Cardiol. 2019;123(4):576–81.

20. Li S, Nemeth I, Donnelly L, Hapca S, Zhou K, Pearson ER. Visit-to-Visit HbA(1c) Variability Is Associated With Cardiovascular Disease and Microvascular Complications in Patients With Newly Diagnosed Type 2 Diabetes. Diabetes Care. 2020;43(2):426–32.

21. Skriver MV, Sandbæk A, Kristensen JK, Støvring H. Relationship of HbA1c variability, absolute changes in HbA1c, and all-cause mortality in type 2 diabetes: a Danish population-based prospective observational study. BMJ Open Diabetes Research & Care. 2015;3(1):e000060.

22. Drozdz D, Alvarez-Pitti J, Wójcik M, Borghi C, Gabbianelli R, Mazur A, et al. Obesity and Cardiometabolic Risk Factors: From Childhood to Adulthood. Nutrients. 2021;13(11).

23. . DSCJ, Dr. Saborni Dey, Dr. Rajesh Ranjan. To determine the correlation between BMI and Glycated Hemoglobin (HbA1c) Level in Patients of Type 2 Diabetes Mellitus. European Journal of Molecular & Clinical Medicine. 2022;9(3):3014–8.

24. Bays HE, Chapman RH, Grandy S. The relationship of body mass index to diabetes mellitus, hypertension and dyslipidaemia: comparison of data from two national surveys. Int J Clin Pract. 2007;61(5):737–47.

