## supplementary_material for "BMI Variability and Cardiovascular Outcomes Within Clinical Trial and Real-World Environments in Type 2 Diabetes: An IMI2 SOPHIA study"

Authors: Robert J Massey^1^, Yu Chen^2^, Marina Panova-Noeva^3^, Michaela Mattheus^3^, Moneeza K Siddiqui^1,4^, Nanette C Schloot^5^, Antonio Ceriello^6^, Ewan R Pearson^1^, Adem Y Dawed^1^

Affiliations: ^1^Population Health & Genomics, University of Dundee, UK; ^2^Lilly Research Laboratories, Eli Lilly and Company, IN, USA; ^3^Boehringer Ingelheim Pharma GmbH & Co. KG, Ingelheim, Germany; ^4^Centre for Primary Care, Wolfson Institute of Population Health, Queen Mary University of London; ^5^Lilly Deutschland GmbH, Bad Homburg, Germany; ^6^IRCCS MultiMedica, Via Milanese 300, 20099 Sesto San Giovanni, MI, Italy.

Appendix 1: Descriptions of the included clinical cohorts

Harmony Outcomes included a total of 9,463 participants with diagnosed type 2 diabetes. These participants were at least 40 years of age with a previous diagnosis of type 2 diabetes and established atherosclerotic cardiovascular disease, and were naïve to GLP-1RA therapy. Patients were randomised to either albiglutide 30 mg/week or placebo on top of standard glycaemic and cardiovascular care. Further details are available in the published trial material (1).

In the REWIND trial, a total of 9,901 participants with diagnosed type 2 diabetes of at least 50 years of age, with HbA1c <9.5%, and BMI >23 kg/m^2^ were included. Participants were randomised to either dulaglutide 1.5 mg/week or placebo. All included participants over 50 years had to have vascular disease; participants over 55 years had to have at least one of the following: documented myocardial ischaemia, or >50% coronary, carotid, or lower extremity artery stenosis, or hypertension with left ventricular hypertrophy, or eGFR <60 mL/min/1·73 m², or albuminuria; and those aged 60 years or above had to have at least two of the following: any tobacco use, use of lipid-controlling therapy or documented dyslipidaemia, use of at least 1 antihypertensive or documented hypertension, or a waist-to-hip ration over 1 in men and 0.8 in women. Further information on key inclusion and exclusion criteria and rationale are available in previously published material (2).

The EMPA-REG OUTCOME trial included a total of 7,028 participants with type 2 diabetes randomised to receive either empagliflozin 10mg/day, empagliflozin 25mg/day, or placebo at a 1:1:1 ratio. These participants were at least 18 years old and had been previously diagnosed with type 2 diabetes. These participants must have had insufficient glycaemic control as defined by HbA1c ≥7.0% and ≤9.0% at screening for anti-diabetic drug naïve participants, or by HbA1c ≥7.0% and ≤10.0% at screening for patients taking background anti-diabetes therapy. These participants must also have been at high risk of cardiovascular events, defined as having at least one of the following: a history of myocardial infarction, single or multi-vessel coronary artery disease, unstable angina, a history of stroke, or occlusive peripheral artery disease. More details on the inclusion and exclusion criteria of this study as well as the rationale can be found in previously published material (3).

Follow-up between the studies was largely similar. Harmony Outcomes required participants to attend in-clinic visits every 4 months throughout the study. The REWIND study required participants to be assessed every 3 to 6 months for the occurrence of cardiovascular and other serious health outcomes, as well as recording of participant HbA1c levels at least every 6 months. The EMPA-REG OUTCOME study required that participants attend follow-up visits every 14 weeks, with an initial period where participants were required to attend visits every 4 weeks up to 16 weeks of follow up, and then a further 36 week period where patients were required to attend visits every 12 weeks. The median follow-up time of Harmony Outcomes was 1.5 years; the median follow-up of REWIND was 5.4 years; and the median observation duration of EMPA-REG OUTCOME was 3.1 years.

The GoDARTS study is sub-study of the DARTS study, which was started in 1996 in order to identify all patients with diabetes within the Tayside region of Scotland using electronic record linkage. GoDARTS began in 1998, when consenting individuals within this electronic database were recruited and invited to provide all phenotypic and genetic data for research purposes. GoSHARE is a similar study occurring across Scotland; participants are asked to allow for their clinical information held within the NHS Scotland electronic medical records to be used for research, as well as for genetic information to be retrieved from blood samples remaining from diagnostic tests. Further descriptions of these databases has been published previously (4, 5).

Appendix 2: The effect of baseline BMI on the association between BMI variability and 3P-MACE

When we stratified individuals in the Harmony Outcomes cohort into either normal weight, overweight, or obese categories, we found no evidence that baseline BMI modified the 3P-MACE risk associated with BMI variability. Individuals of a normal weight (n = 660) experienced no significant increase in 3P-MACE risk per +1 SD increase in ASV BMI in model 5(HR 1.31, 95% CI 0.69 – 2.49, P = 0.40; Table S5). In overweight individuals (n = 2753), a significantly increased risk of 3P-MACE was associated with a +1 SD increase in ASV BMI (HR 1.74, 95% CI 1.29 – 2.35, P < 0.001; Table S5). Similarly, in individuals with obesity (n = 5534), a +1 SD increase in ASV BMI was associated with an increase in 3P-MACE risk (HR 1.45, 95% CI 1.18 – 1.78, P < 0.001; Table S5).

When we performed this analysis in the REWIND cohort, we similarly found that baseline BMI did not modify the association between BMI variability and 3P-MACE risk. In individuals of normal weight (n = 287), an increase of ASV BMI by +1 SD was not significantly associated with an increased risk of 3P-MACE (HR 1.28, 95% CI 0.96 – 1.70, P = 0.095; Figure S4(a)). In overweight participants (n = 1383), no significant increase in 3P-MACE risk was again observed when ASV BMI increased by +1 SD (HR 1.12, 95% CI 0.96 – 1.30, P = 0.154; Figure S5(a)). In participants with obesity (n = 2770), no significant association between 3P-MACE and an increase in ASV BMI by +1 SD was again observed (HR 1.08, 95% CI 0.99 – 1.17, P = 0.075; Figure S6(a)).

When we performed our analysis within the EMPA-REG OUTCOME cohort, we again found little evidence of a relationship between baseline BMI and 3P-MACE risk associated with BMI variability. Participants with a normal weight (n = 282) experienced no significant change in 3P-MACE risk when ASV BMI increased by +1 SD (HR 0.60, 95% CI 0.28 – 1.26, P = 0.1749; Table S5). A similar result was found within overweight participants (n = 767); an increase in ASV BMI by +1 SD was not associated with a significant change in 3P-MACE risk (HR 0.94, 95% CI 0.67 – 1.32, P = 0.7309; Table S7). In obese participants (n = 1133), an increase in ASV BMI by +1 SD was observed to be associated with a decrease in 3P-MACE risk (HR 0.72, 95% CI 0.53 – 0.96, P = 0.0276; Table S8).

Finally, within the Tayside Bioresource cohort, we observed no obvious effect modification evidence of baseline BMI on the association between BMI variability and 3P-MACE risk. A +1 SD increase in ASV BMI within individuals with normal weight (n = 787) was significantly associated with an increase in 3P-MACE risk (HR 1.38, 95% CI 1.11 – 1.70, P = 0.003; Figure S4(b)). A similar result was observed within overweight participants (n = 2287), where a +1 SD increase in ASV BMI was associated with significantly increased 3P-MACE risk (HR 1.27, 95% CI 1.08 – 1.50, P = 0.005; Figure S5(b)). Within obese participants (n = 3580), a slightly smaller but still significantly increased risk of 3P-MACE was associated with a +1 SD increase in ASV BMI (HR 1.10, 95% CI 1.04 – 1.20, P = 0.002; Figure S6(b)).

### Clinical Features

Table S1: A table of the clinical features of the participants of the Harmony Outcomes (n = 9198), REWIND (n = 4440), EMPA-REG OUTCOME (n = 2333), and Tayside Bioresource (n = 6980) cohorts. Data within round brackets refers to standard deviation; data within square brackets refers to range. ⁋ = includes current and former smokers; † = data from a smaller subset, n = 6,654; ‡ = value is ASVT, as stated in methodology; * = minimum of 3 measurements; CVD = cardiovascular disease.

| **Clinical Feature** | **Harmony Outcomes n=9198** | **REWIND n=4440** | **EMPA-REG OUTCOME n=2333** | **Tayside Bioresource n=6980** |
| --- | --- | --- | --- | --- |
| **Age (years)** | 65 (9.0) | 66.0 (6.5) | 63.2 (8.8) | 59.6 (11.0) |
| **Male (%)** | 69.5 | 53.6 | 72.0 | 56.7 |
| **Current smokers (%)** | 15.1 | 14.4 | 13.9 | 44.3⁋ |
| **BMI (kg/m^2^)** | 32.3 (5.9) | 32.4 (5.7) | 30.7 (5.2) | 31.3 (6.1) |
| **History of CVD (%)** | NA | 18.2 | 75.6 | NA |
| **Systolic BP (mmHg)** | 134.8 (16.5) | 139 (18.4) | 135.8 (17.2) | NA |
| **T2D duration (years)** | 14.1 (8.7) | 10.6 (7.2) | <=1, n = 52;  >1 to <=5, n = 371;  >5 to <=10, n = 571;  >10, n = 1339 | 7 (3.9) |
| **LDL-C (mg/dL)** | NA | 98.5 (37.5) | 84.9 (35.3) | NA |
| **HDL-C (mg/dL)** | NA | 45.8 (13.8) | 44.0 (11.3) | 49.1 (13.7)† |
| **Total cholesterol (mg/dL)** | NA | 175 (44.9) | 161.9 (43.1) | 181.0 (41.6)† |
| **HbA1c (%)** | 8.7 (1.5) | 7.3 (1.1) | 8.1 (0.8) | 7.5 (1.4) |
| **Num. of BMI (n)** | 4 [3-14] | 6.5 (1.3) | NA* | 8.4 (2.6) |
| **ASV BMI** | 0.57 (0.5) | 0.97 (0.6) | 0.74 (0.7) | 0.0037 (0.0024)‡ |
| **ASV HbA1c** | 0.66 (0.5) | 0.75 (0.5) | 0.66 (0.4) | 0.064 (0.1)‡ |

### Comparing Quartiles of BMI variability

Table S2**:** *A table summarising the adjusted hazard ratio (HR) and 95% confidence interval (CI) of 3-point major adverse cardiovascular events (MACE) risk for individuals within quartile 2, quartile 3, and quartile 4 (using quartile 1 as reference) of BMI variability within the EMPA-REG OUTCOME (n = 2333) trial placebo-arm cohort.* Risk estimates presented based on Cox regression model.

| **Covariate** | **HR** | **95% CI** | **P value** | **Overall P value** |
| --- | --- | --- | --- | --- |
| Age | 1.01 | 1.00 - 1.03 | | 0.1385 |
| Baseline BMI | 0.95 | 0.77 - 1.13 | | 0.5512 |
| Baseline HbA1c | 0.84 | 0.70 - 1.00 | | 0.0547 |
| Average BMI | 1.04 | 0.87 - 1.23 | | 0.6931 |
| Number of BMI measures | 0.25 | 0.22 - 0.29 | | <0.0001 |
| Baseline SBP | 1.01 | 1.00 - 1.01 | | 0.0857 |
| Lipid-lowering drug use | | |  |  |
| No (REF) | 1 |  |  | 0.1252 |
| Yes | 1.33 | 0.93 - 1.91 | 0.1252 |  |
| Quartiles of BMI ASV | |  |  |  |
| 1 (REF) | 1 |  |  | 0.0727 |
| 2 | 0.88 | 0.61 - 1.28 | 0.51 |  |
| 3 | 0.74 | 0.49 - 1.11 | 0.1457 |  |
| 4 | 0.61 | 0.42 - 0.90 | 0.0115 |  |
| T2D duration | |  |  |  |
| <= 1yr (REF) | 1 |  |  | 0.6347 |
| <= 5yrs but >1yr | 2.56 | 0.6 - 10.97 | 0.2049 |  |
| <= 10rys but >5yrs | 2.5 | 0.59 - 10.55 | 0.2111 |  |
| >10 yrs | 2.57 | 0.62 - 10.56 | 0.1924 |  |
| Gender |  |  |  |  |
| Male (REF) | 1 |  |  | 0.3882 |
| Female | 0.86 | 0.61 - 1.21 | 0.3882 |  |
| Smoking status | |  |  |  |
| Never smoked (REF) | 1 |  |  | 0.2584 |
| Ex-smoker | 0.8 | 0.58 - 1.12 | 0.2003 |  |
| Currently smokes | 1.08 | 0.70 - 1.67 | 0.7365 |  |

Figure S1(a)**:** *A forest plot summarising the adjusted hazard ratio (HR) and 95% confidence interval (CI) of 3-point major adverse cardiovascular events (MACE) risk for individuals within quartile 2, quartile 3, and quartile 4 (using quartile 1 as reference) of BMI variability within the Harmony Outcomes (n = 9198)* *trial cohort after adjustment for treatment, baseline BMI, sex, age, smoking, type 2 diabetes duration, systolic blood pressure, statin use, baseline HbA1c, and number of BMI measurement. Risk estimates for each covariate are calculated via multivariate Cox regression.*

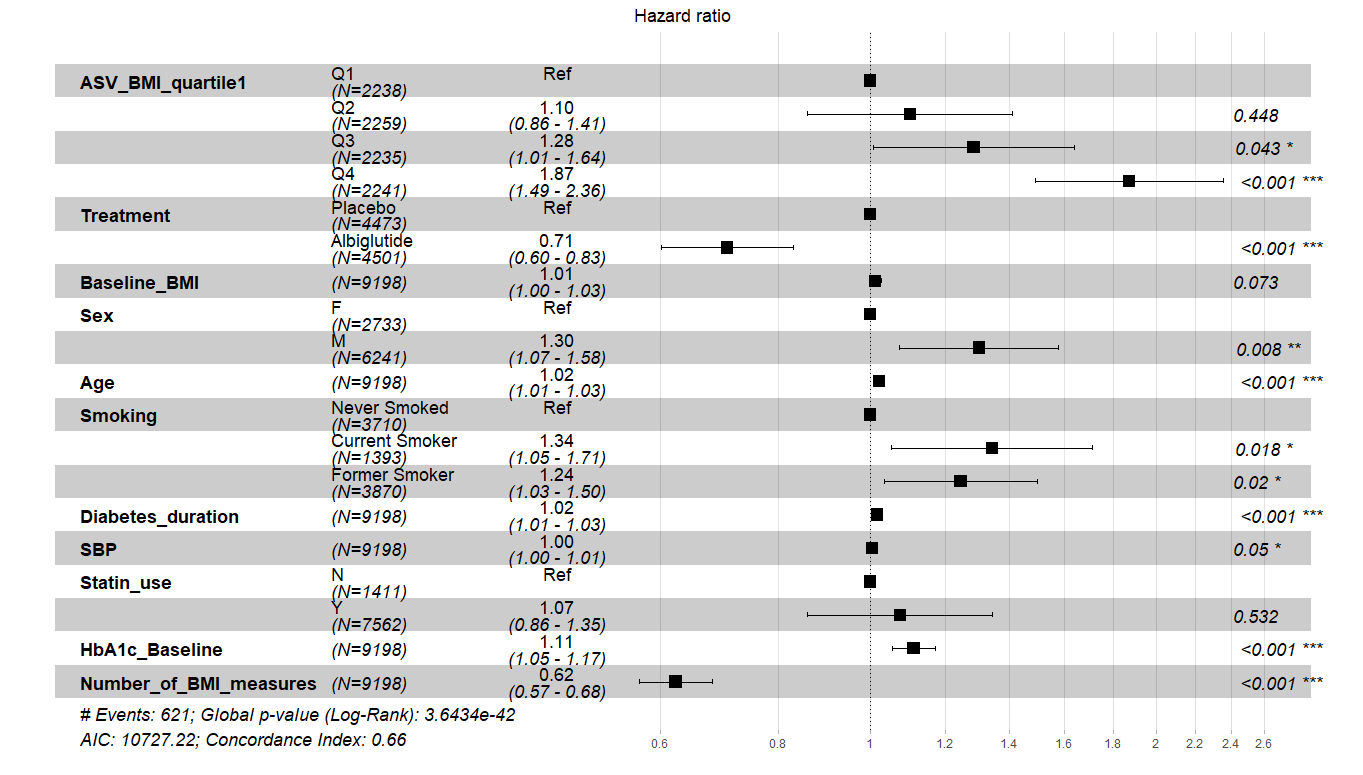

Figure S1(b)**:** *A forest plot summarising the adjusted hazard ratio (HR) and 95% confidence interval (CI) of 3-point major adverse cardiovascular events (MACE) risk for individuals within quartile 2, quartile 3, and quartile 4 (using quartile 1 as reference) of BMI variability within the REWIND (n = 4440)* *trial placebo-arm cohort after adjustment for age, sex, baseline BMI, systolic blood pressure, baseline HbA1c, history of coronary artery disease, statin use, smoking, and number of BMI measurement. Risk estimates for each covariate are calculated via multivariate Cox regression.*

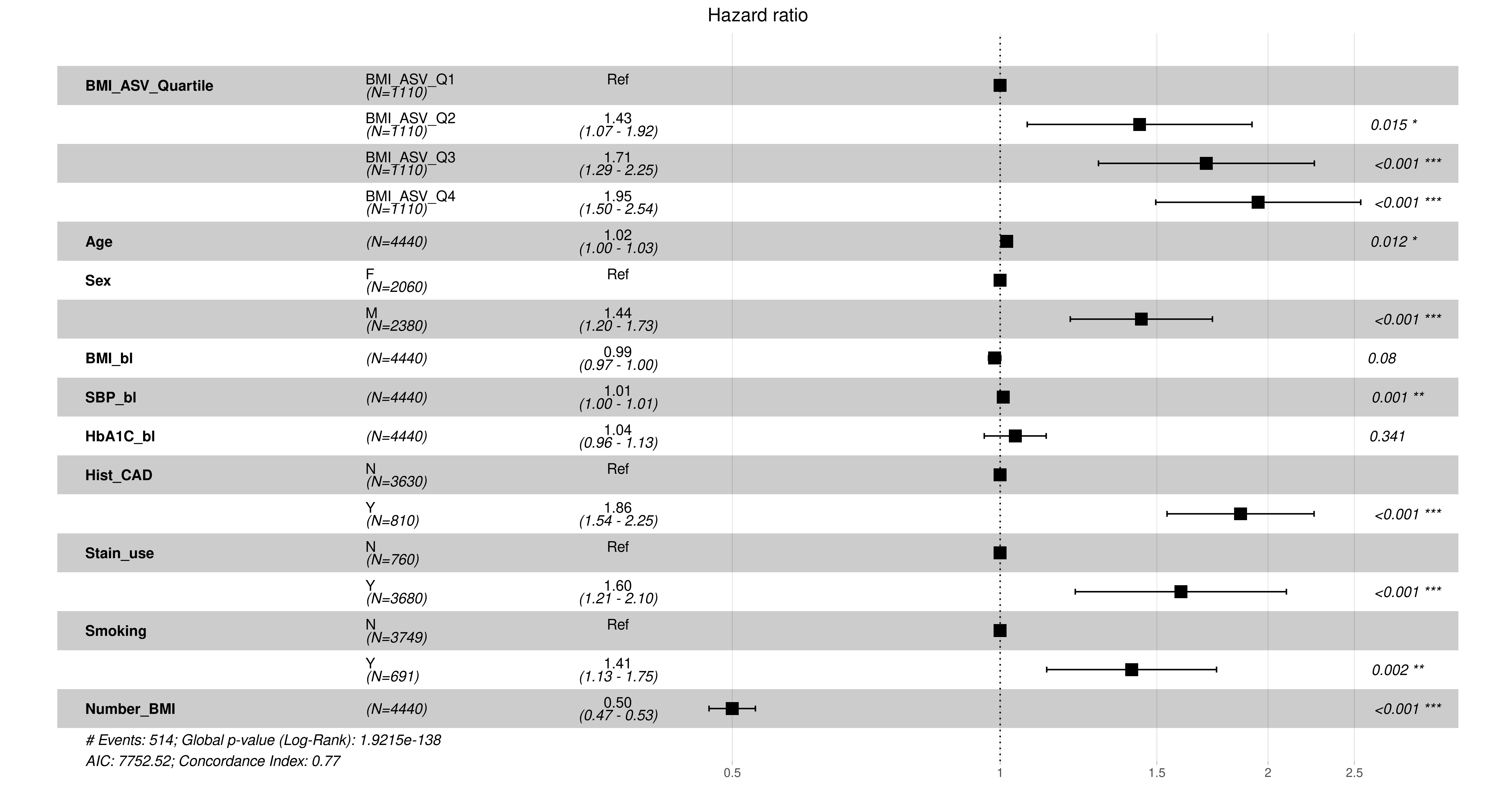

Figure S1(c)**:** *A forest plot summarising the adjusted hazard ratio (HR) and 95% confidence interval (CI) of 3-point major adverse cardiovascular events (MACE) risk for individuals within quartile 2, quartile 3, and quartile 4 (using quartile 1 as reference) of BMI variability within the Tayside Bioresource (n = 6980)* *cohort after adjustment for age, sex, baseline BMI, systolic blood pressure, baseline HbA1c, history of coronary artery disease, statin use, smoking, and number of BMI measurement. The covariate “age” has been split into quartiles for this model in order to allow the model to meet proportional hazards assumptions. Risk estimates for each covariate are calculated via multivariate Cox regression.*

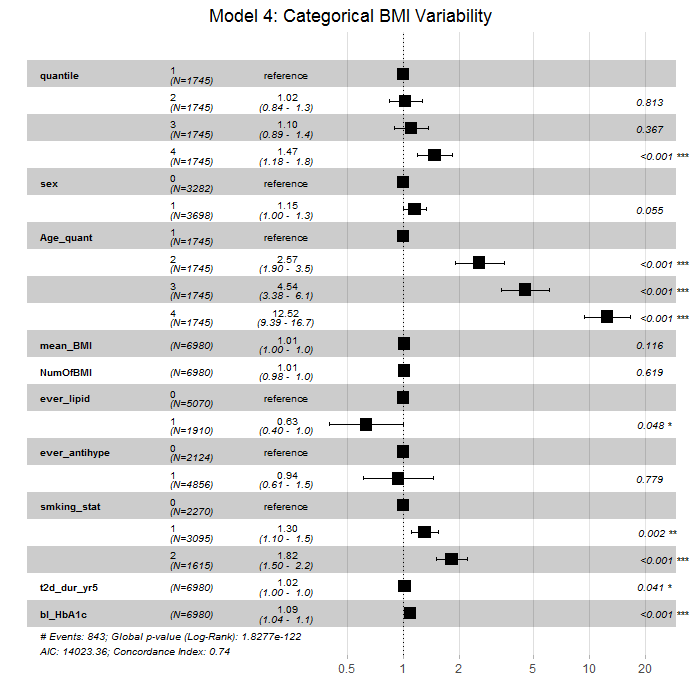

### Models including HbA1c variability

Table S3**:** *A table summarising the adjusted hazard ratio (HR) and 95% confidence interval (CI) of 3-point major adverse cardiovascular events (MACE) risk associated with a +1 standard deviation (SD) increase in BMI variability within the EMPA-REG OUTCOME (n = 2333) trial placebo-arm cohort.* Risk estimates presented based on Cox regression model.

| **Covariate** | **HR** | **95% CI** | **P value** | **Overall P value** |
| --- | --- | --- | --- | --- |
| Age | 1.01 | 1.00 - 1.03 | | 0.1552 |
| Baseline BMI | 0.94 | 0.78 - 1.14 | | 0.5551 |
| Baseline HbA1c | 0.82 | 0.68 - 0.98 | | 0.0333 |
| Average BMI | 1.04 | 0.86 - 1.27 | | 0.6569 |
| Number of BMI measures | 0.24 | 0.21 - 0.28 | | <0.0001 |
| BMI ASV | 0.8 | 0.66 - 0.97 | | 0.0261 |
| Baseline SBP | 1.01 | 1.00 - 1.01 | | 0.167 |
| HbA1c ASV | 1.04 | 0.93 - 1.17 | | 0.4764 |
| Lipid-lowering drug use |  |  |  |  |
| No (REF) | 1 |  |  | 0.1242 |
| Yes | 1.34 | 0.92 - 1.93 | 0.1242 |  |
| T2D duration |  |  |  |  |
| <= 1yr (REF) | 1 |  |  | 0.522 |
| <= 5yrs but >1yr | 2.71 | 0.63 - 11.60 | 0.1786 |  |
| <= 10rys but >5yrs | 2.87 | 0.68 - 12.10 | 0.1521 |  |
| >10 yrs | 2.82 | 0.68 - 11.68 | 0.1525 |  |
| Gender |  |  |  |  |
| Male (REF) | 1 |  |  | 0.4696 |
| Female | 0.88 | 0.62 - 1.24 | 0.4696 |  |
| Smoking status |  |  |  |  |
| Never smoked (REF) | 1 |  |  | 0.2882 |
| Ex-smoker | 0.81 | 0.58 - 1.14 | 0.225 |  |
| Currently smokes | 1.08 | 0.70 - 1.69 | 0.7198 |  |

Table S4**:** *A table summarising the adjusted hazard ratio (HR) and 95% confidence interval (CI) of 3-point major adverse cardiovascular events (MACE) risk for individuals within quartile 2, quartile 3, and quartile 4 (using quartile 1 as reference) of BMI variability within the EMPA-REG OUTCOME (n = 2333) trial placebo-arm cohort. ASV HbA1c variability is included as an additional covariate.* Risk estimates presented based on Cox regression model.

| **Covariate** | **HR** | **95% CI** | **P value** | **Overall P value** |
| --- | --- | --- | --- | --- |
| Age | 1.01 | 1.00 - 1.03 | | 0.1336 |
| Baseline BMI | 0.95 | 0.81 - 1.13 | | 0.59 |
| Baseline HbA1c | 0.82 | 0.68 - 0.99 | | 0.0381 |
| Average BMI | 1.03 | 0.87 - 1.22 | | 0.711 |
| Number of BMI measures | 0.25 | 0.21- 0.28 | | <0.0001 |
| Baseline SBP | 1.01 | 1.00 - 1.01 | | 0.1654 |
| HbA1c ASV | 1.04 | 0.93 - 1.17 | | 0.4871 |
| Lipid-lowering drug use |  |  |  |  |
| No (REF) | 1 |  |  | 0.0994 |
| Yes | 1.36 | 0.94 - 1.97 | 0.994 |  |
| Quartiles of BMI ASV |  |  |  |  |
| 1 (REF) | 1 |  |  | 0.0995 |
| 2 | 0.91 | 0.62 - 1.32 | 0.6127 |  |
| 3 | 0.73 | 0.48 - 1.09 | 0.1215 |  |
| 4 | 0.64 | 0.44 - 0.93 | 0.0212 |  |
| T2D duration |  |  |  |  |
| <= 1yr (REF) | 1 |  |  | 0.6164 |
| <= 5yrs but >1yr | 2.55 | 0.60 - 10.95 | 0.2066 |  |
| <= 10rys but >5yrs | 2.66 | 0.63 - 11.23 | 0.1842 |  |
| >10 yrs | 2.62 | 0.63 - 10.86 | 0.1847 |  |
| Gender |  |  |  |  |
| Male (REF) | 1 |  |  | 0.4701 |
| Female | 0.88 | 0.62 - 1.24 | 0.4701 |  |
| Smoking status |  |  |  |  |
| Never smoked (REF) | 1 |  |  | 0.2974 |
| Ex-smoker | 0.82 | 0.58 - 1.14 | 0.2313 |  |
| Currently smokes | 1.08 | 0.69 - 1.68 | 0.7296 |  |

Figure S2(a)**:** *A forest plot summarising the adjusted hazard ratio (HR) and 95% confidence interval (CI) of 3-point major adverse cardiovascular events (MACE) risk associated with a +1 standard deviation (SD) increase in BMI variability within the Harmony Outcomes (n = 9198) trial cohort after adjustment for treatment, baseline BMI, sex, age, smoking, type 2 diabetes duration, systolic blood pressure, statin use, baseline HbA1c, and number of BMI measurement. Risk estimates for each covariate are calculated via multivariate Cox regression. ASV HbA1c variability is included as an additional covariate.*

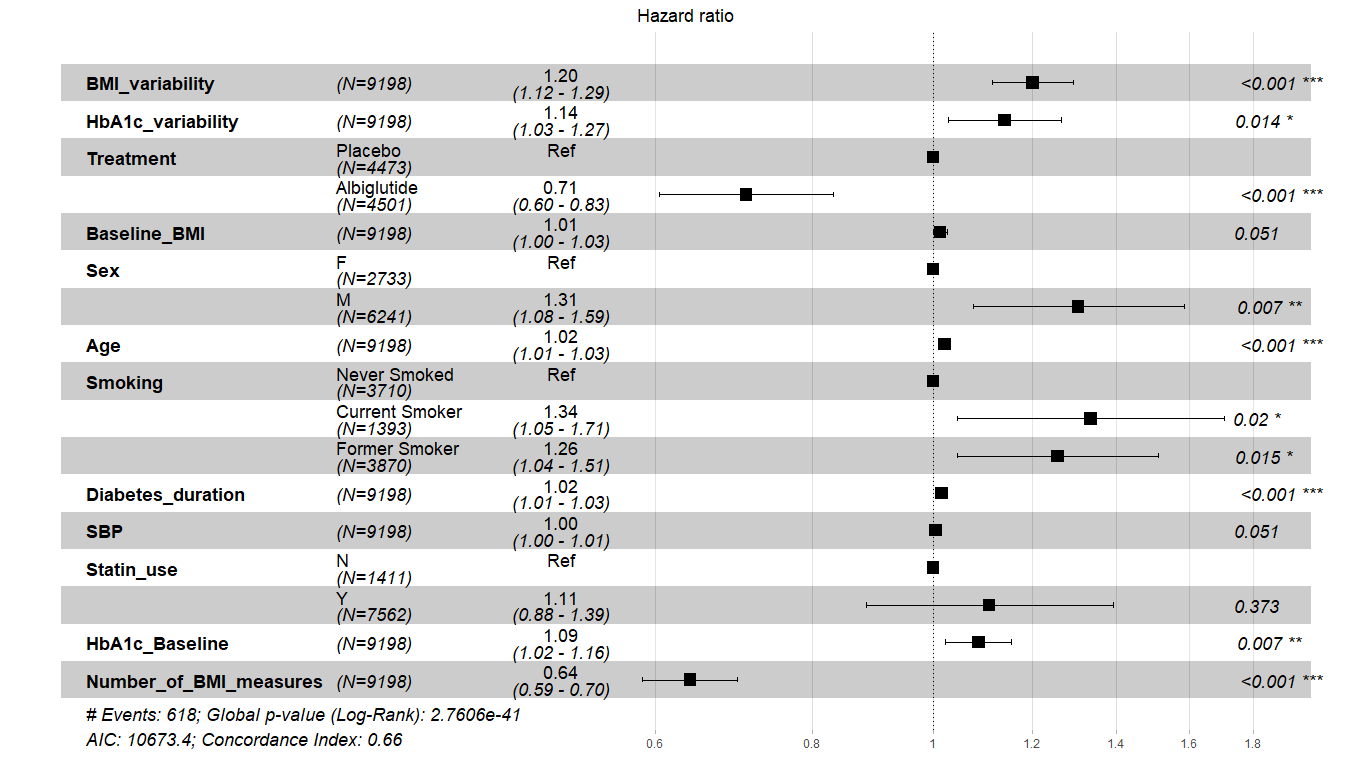

Figure S2(b)**:** *A forest plot summarising the adjusted hazard ratio (HR) and 95% confidence interval (CI) of 3-point major adverse cardiovascular events (MACE) risk associated with a +1 standard deviation (SD) increase in BMI variability within the REWIND (n = 4440) trial placebo-arm cohort after adjustment for age, sex, baseline BMI, systolic blood pressure, baseline HbA1c, history of coronary artery disease, statin use, smoking, and number of BMI measurement. Risk estimates for each covariate are calculated via multivariate Cox regression. ASV HbA1c variability is included as an additional covariate.*

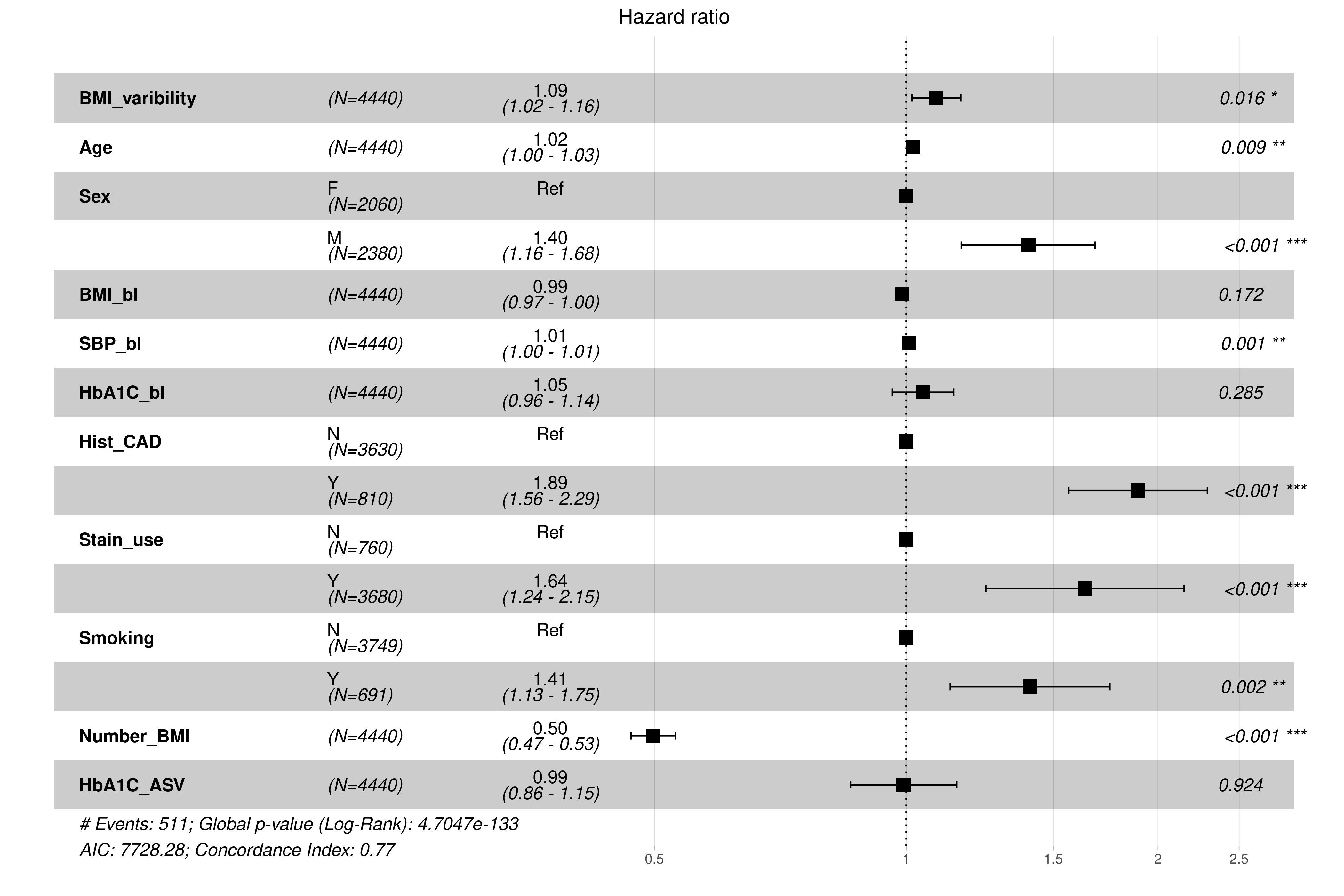

Figure S2(c)**:** *A forest plot summarising the adjusted hazard ratio (HR) and 95% confidence interval (CI) of 3-point major adverse cardiovascular events (MACE) risk associated with a +1 standard deviation (SD) increase in BMI variability within the Tayside Bioresource (n = 6980) cohort. after adjustment for age, sex, baseline BMI, systolic blood pressure, baseline HbA1c, history of coronary artery disease, statin use, smoking, and number of BMI measurement. The covariate “age” has been split into quartiles for this model in order to allow the model to meet proportional hazards assumptions. Risk estimates for each covariate are calculated via multivariate Cox regression. ASV HbA1c variability is included as an additional covariate.*

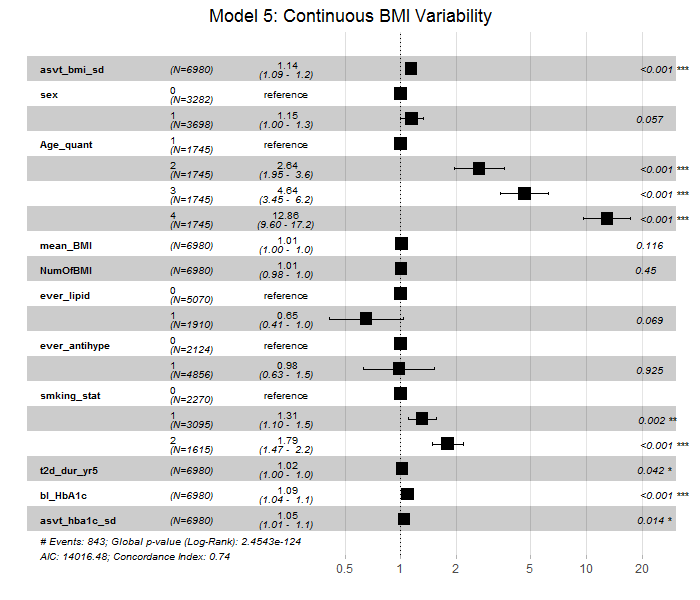

Figure S3(a)**:** *A forest plot summarising the adjusted hazard ratio (HR) and 95% confidence interval (CI) of 3-point major adverse cardiovascular events (MACE) risk for individuals within quartile 2, quartile 3, and quartile 4 (using quartile 1 as reference) of BMI variability within the Harmony Outcomes* (n = 9198) *trial cohort after adjustment for treatment, baseline BMI, sex, age, smoking, type 2 diabetes duration, systolic blood pressure, statin use, baseline HbA1c, and number of BMI measurement. Risk estimates for each covariate are calculated via multivariate Cox regression. ASV HbA1c variability is included as an additional covariate.*

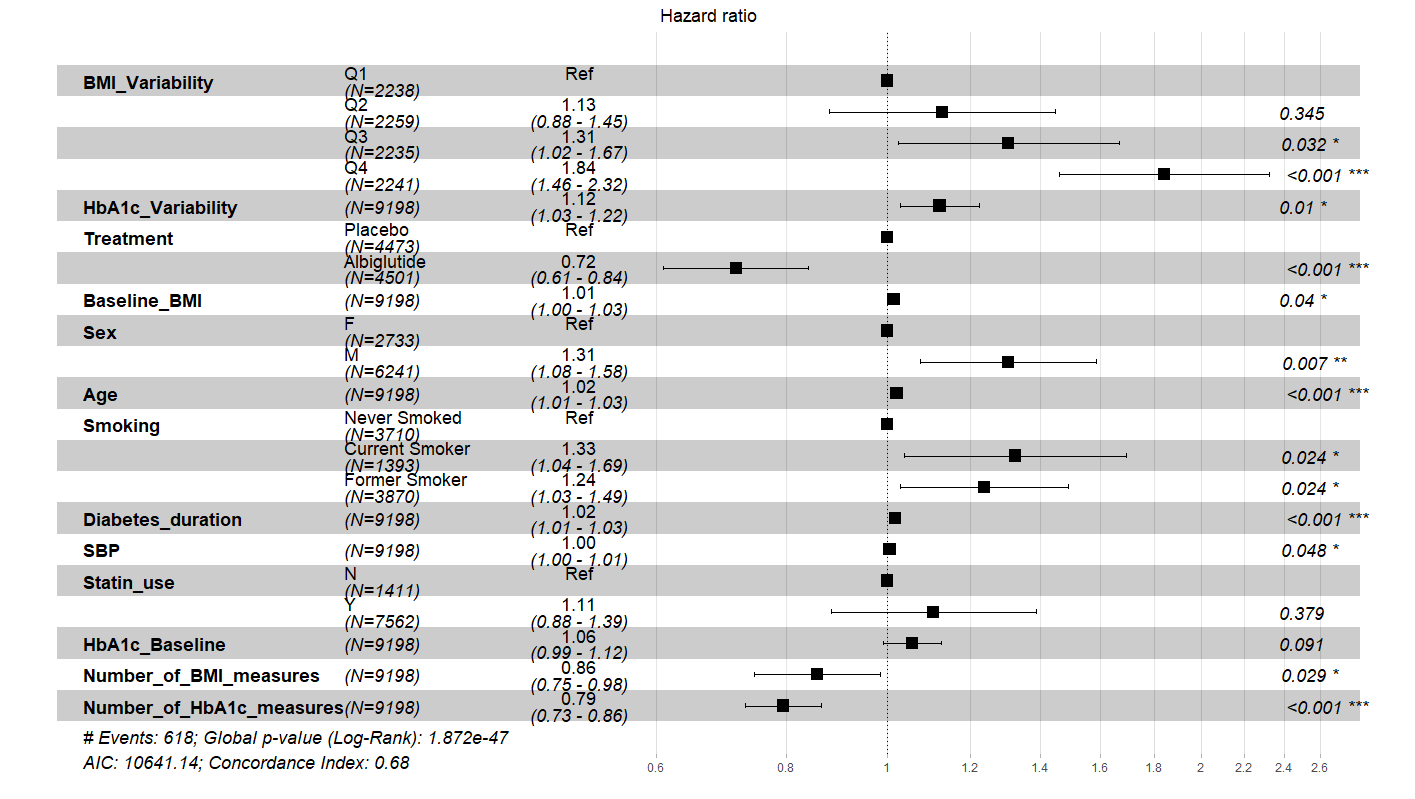

Figure S3(b)**:** *A forest plot summarising the adjusted hazard ratio (HR) and 95% confidence interval (CI) of 3-point major adverse cardiovascular events (MACE) risk for individuals within quartile 2, quartile 3, and quartile 4 (using quartile 1 as reference) of BMI variability within the REWIND (n = 4440) trial placebo-arm cohort after adjustment for age, sex, baseline BMI, systolic blood pressure, baseline HbA1c, history of coronary artery disease, statin use, smoking, and number of BMI measurement. Risk estimates for each covariate are calculated via multivariate Cox regression. ASV HbA1c variability is included as an additional covariate.*

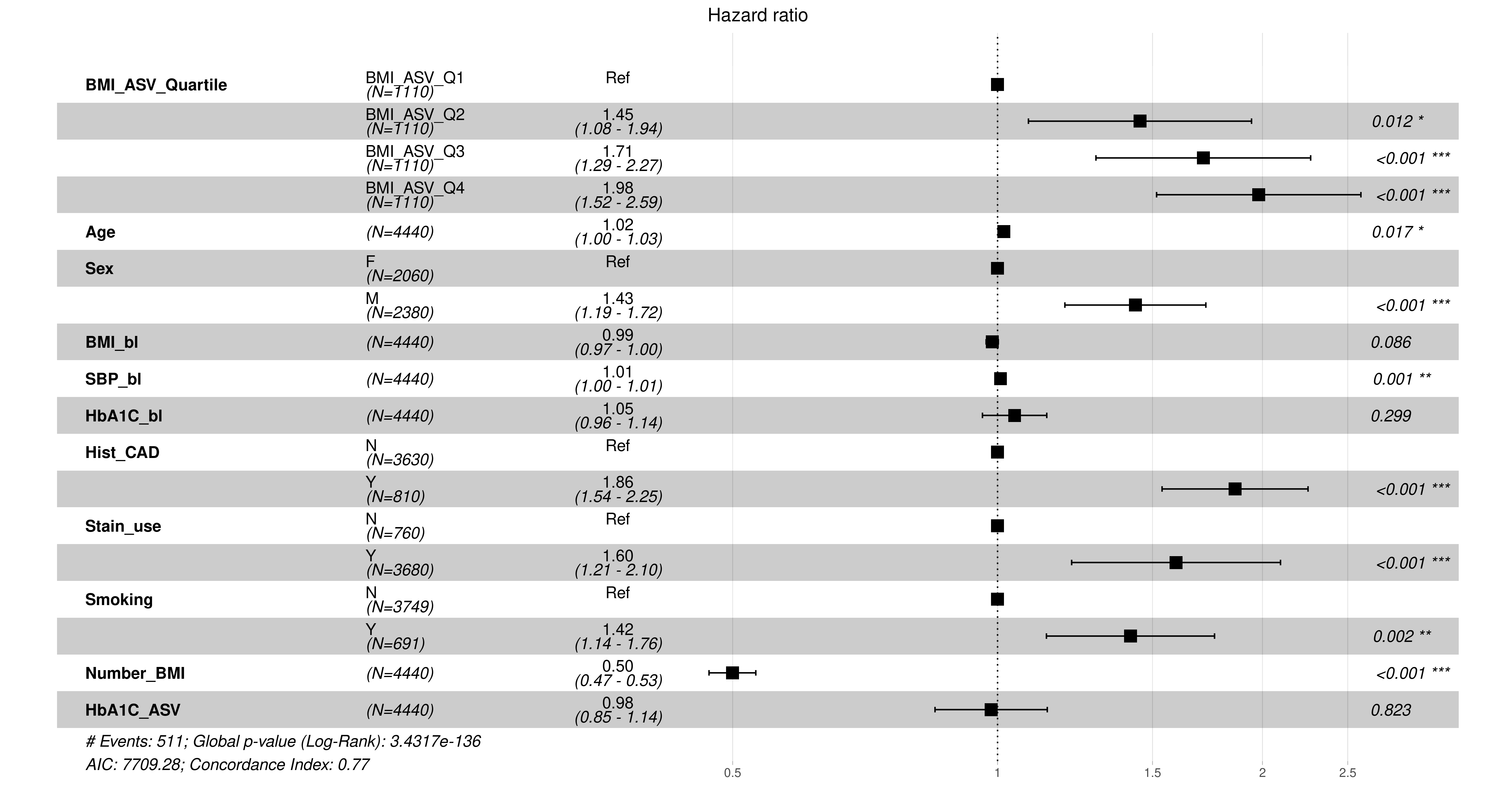

Figure S3(c)**:** *A forest plot summarising the adjusted hazard ratio (HR) and 95% confidence interval (CI) of 3-point major adverse cardiovascular events (MACE) risk for individuals within quartile 2, quartile 3, and quartile 4 (using quartile 1 as reference) of BMI variability within the Tayside Bioresource (n = 6980)* *cohort after adjustment for age, sex, baseline BMI, systolic blood pressure, baseline HbA1c, history of coronary artery disease, statin use, smoking, and number of BMI measurement. The covariate “age” has been split into quartiles for this model in order to allow the model to meet proportional hazards assumptions. Risk estimates for each covariate are calculated via multivariate Cox regression. ASV HbA1c variability is included as an additional covariate.*

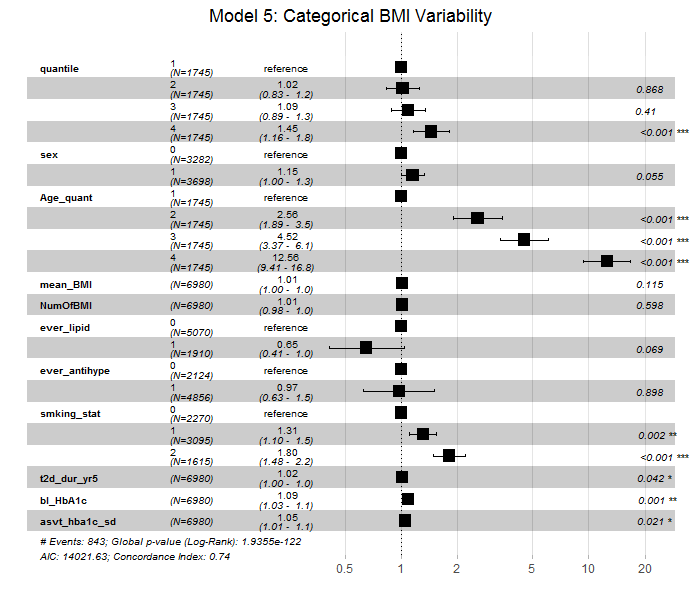

### Baseline BMI and BMI Variability

Table S5**:** *A table summarising the adjusted hazard ratio (HR) and 95% confidence interval (CI) of 3-point major adverse cardiovascular events (MACE) risk associated with a +1 standard deviation (SD) increase in BMI variability within the Harmony Outcomes (n = 9198)* *trial cohort when stratified into normal, overweight, or obese BMI at baseline. ASV HbA1c variability is included as an additional covariate.* The fully adjusted model is adjusted for treatment, baseline BMI, sex, age, smoking, type 2 diabetes duration, systolic blood pressure, statin use, baseline HbA1c, and number of BMI measurement. Risk estimates are calculated via Cox regression.

| **Outcome** | **Unadjusted HR [95% CIs]** | **P** | **Fully adjusted HR [95% CIs]** | **P** |
| --- | --- | --- | --- | --- |
| Normal weight | 1.14[0.62-2.08] | 0.68 | 1.31[0.69-2.49] | 0.40 |
| Overweight | 1.74[1.18-2.13] | 0.002 | 1.74[2.29-2.35] | 0.0003 |
| Obese | 1.31[1.07-1.61] | 0.009 | 1.45[1.18-1.78] | 0.0004 |

Table S6**:** *A table summarising the adjusted hazard ratio (HR) and 95% confidence interval (CI) of 3-point major adverse cardiovascular events (MACE) risk associated with a +1 standard deviation (SD) increase in BMI variability within the EMPA-REG OUTCOME (n = 2333) trial placebo-arm cohort in individuals with normal baseline BMI.* Risk estimates presented based on Cox regression model.

| **Covariate** | **HR** | **95% CI** | **P value** | **Overall P value** |
| --- | --- | --- | --- | --- |
| Age | 0.99 | 0.94 - 1.03 | | 0.5368 |
| Baseline BMI | 0.65 | 0.33 - 1.31 | | 0.2293 |
| Baseline HbA1c | 0.9 | 0.51 - 1.58 | | 0.7174 |
| Average BMI | 1.19 | 0.51 - 1.58 | | 0.5951 |
| Number of BMI measures | 0.22 | 0.62 - 2.30 | | <0.0001 |
| BMI ASV | 0.72 | 0.14 - 0.36 | | 0.3854 |
| Baseline SBP | 1.01 | 0.34 - 1.52 | | 0.4831 |
| HbA1c ASV | 1.17 | 0.98 - 1.03 | | 0.3686 |
| Lipid-lowering drug use |  |  |  |  |
| No (REF) | 1 |  |  | 0.3022 |
| Yes | 1.97 | 0.54 - 7.15 | 0.3022 |  |
| T2D duration |  |  |  |  |
| <= 1yr (REF) | 1 |  |  | 0.4308 |
| <= 5yrs but >1yr | 1 |  |  |  |
| <= 10rys but >5yrs | 1 |  |  |  |
| >10 yrs | 1 |  |  |  |
| Gender |  |  |  |  |
| Male (REF) | 1 |  |  | 0.1253 |
| Female | 0.38 | 0.11 - 1.31 | 0.1253 |  |
| Smoking status |  |  |  |  |
| Never smoked (REF) | 1 |  |  | 0.6975 |
| Ex-smoker | 0.64 | 0.20 - 2.05 | 0.4556 |  |
| Currently smokes | 0.61 | 0.15 - 2.53 | 0.4992 |  |

Table S7**:** *A table summarising the adjusted hazard ratio (HR) and 95% confidence interval (CI) of 3-point major adverse cardiovascular events (MACE) risk associated with a +1 standard deviation (SD) increase in BMI variability within EMPA-REG OUTCOME (n = 2333) trial placebo-arm cohort in individuals with overweight baseline BMI.* Risk estimates presented based on Cox regression model.

| **Covariate** | **HR** | **95% CI** | **P value** | **Overall P value** |
| --- | --- | --- | --- | --- |
| Age | 1.01 | 0.99 - 1.04 | | 0.2787 |
| Baseline BMI | 0.82 | 0.57 - 1.16 | | 0.2631 |
| Baseline HbA1c | 0.92 | 0.70 - 1.21 | | 0.5598 |
| Average BMI | 1.04 | 0.75 - 1.43 | | 0.8162 |
| Number of BMI measures | 0.2 | 0.15 - 0.26 | | <0.0001 |
| BMI ASV | 0.94 | 0.67 - 1.31 | | 0.7015 |
| Baseline SBP | 0.99 | 0.98 - 1.01 | | 0.3194 |
| HbA1c ASV | 1.03 | 0.87 - 1.21 | | 0.7533 |
| Lipid-lowering drug use |  |  |  |  |
| No (REF) | 1 |  |  | 0.6606 |
| Yes | 1.16 | 0.60 - 2.27 | 0.6606 |  |
| T2D duration |  |  |  |  |
| <= 1yr (REF) | 1 |  |  | 0.0086 |
| <= 5yrs but >1yr | 0.97 | 0.11 - 8.97 | 0.9821 |  |
| <= 10rys but >5yrs | 3.27 | 0.40 - 26.89 | 0.2702 |  |
| >10 yrs | 4.13 | 0.52 - 32.74 | 0.1791 |  |
| Gender |  |  |  |  |
| Male (REF) | 1 |  |  | 0.4147 |
| Female | 1.27 | 0.71 - 2.26 | 0.4147 |  |
| Smoking status |  |  |  |  |
| Never smoked (REF) | 1 |  |  | 0.0432 |
| Ex-smoker | 0.73 | 0.42 - 1.29 | 0.2784 |  |
| Currently smokes | 1.58 | 0.82 - 3.03 | 0.1725 |  |

Table S8**:** *A table summarising the adjusted hazard ratio (HR) and 95% confidence interval (CI) of 3-point major adverse cardiovascular events (MACE) risk associated with a +1 standard deviation (SD) increase in BMI variability within the EMPA-REG OUTCOME (n = 2333) trial placebo-arm cohort in individuals with obese baseline BMI.* Risk estimates presented based on Cox regression model.

| **Covariate** | **HR** | **95% CI** | **P value** | **Overall P value** |
| --- | --- | --- | --- | --- |
| Age | 1.03 | 1.00 - 1.07 | | 0.025 |
| Baseline BMI | 1.01 | 0.75 - 1.35 | | 0.9648 |
| Baseline HbA1c | 0.75 | 0.56 - 0.99 | | 0.0448 |
| Average BMI | 1 | 0.75 - 1.32 | | 0.9828 |
| Number of BMI measures | 0.23 | 0.19 - 0.29 | | <0.0001 |
| BMI ASV | 0.71 | 0.53 - 0.95 | | 0.0231 |
| Baseline SBP | 1.01 | 1.00 - 1.02 | | 0.1082 |
| HbA1c ASV | 1.16 | 0.93 - 1.45 | | 0.1828 |
| Lipid-lowering drug use |  |  |  |  |
| No (REF) | 1 |  |  | 0.7923 |
| Yes | 1.07 | 0.64 - 1.81 | 0.7923 |  |
| T2D duration |  |  |  |  |
| <= 1yr (REF) | 1 |  |  | 0.1052 |
| <= 5yrs but >1yr | 3.11 | 0.40 - 24.34 | 0.2797 |  |
| <= 10rys but >5yrs | 1.4 | 0.18 - 11.19 | 0.7495 |  |
| >10 yrs | 1.81 | 0.24 - 13.79 | 0.5678 |  |
| Gender |  |  |  |  |
| Male (REF) | 1 |  |  | 0.2419 |
| Female | 0.74 | 0.44 - 1.23 | 0.2419 |  |
| Smoking status |  |  |  |  |
| Never smoked (REF) | 1 |  |  | 0.7552 |
| Ex-smoker | 0.84 | 0.52 - 1.36 | 0.4874 |  |
| Currently smokes | 0.81 | 0.39 - 1.71 | 0.584 |  |

Figure S4(a)**:** *A forest plot summarising the adjusted hazard ratio (HR) and 95% confidence interval (CI) of 3-point major adverse cardiovascular events (MACE) risk associated with a +1 standard deviation (SD) increase in BMI variability within REWIND (n = 4440) trial placebo-arm cohort in individuals with normal baseline BMI after adjustment for treatment, baseline BMI, sex, age, smoking, type 2 diabetes duration, systolic blood pressure, statin use, baseline HbA1c, number of BMI measurement, and ASV HbA1c. Risk estimates for each covariate are calculated via multivariate Cox regression.*

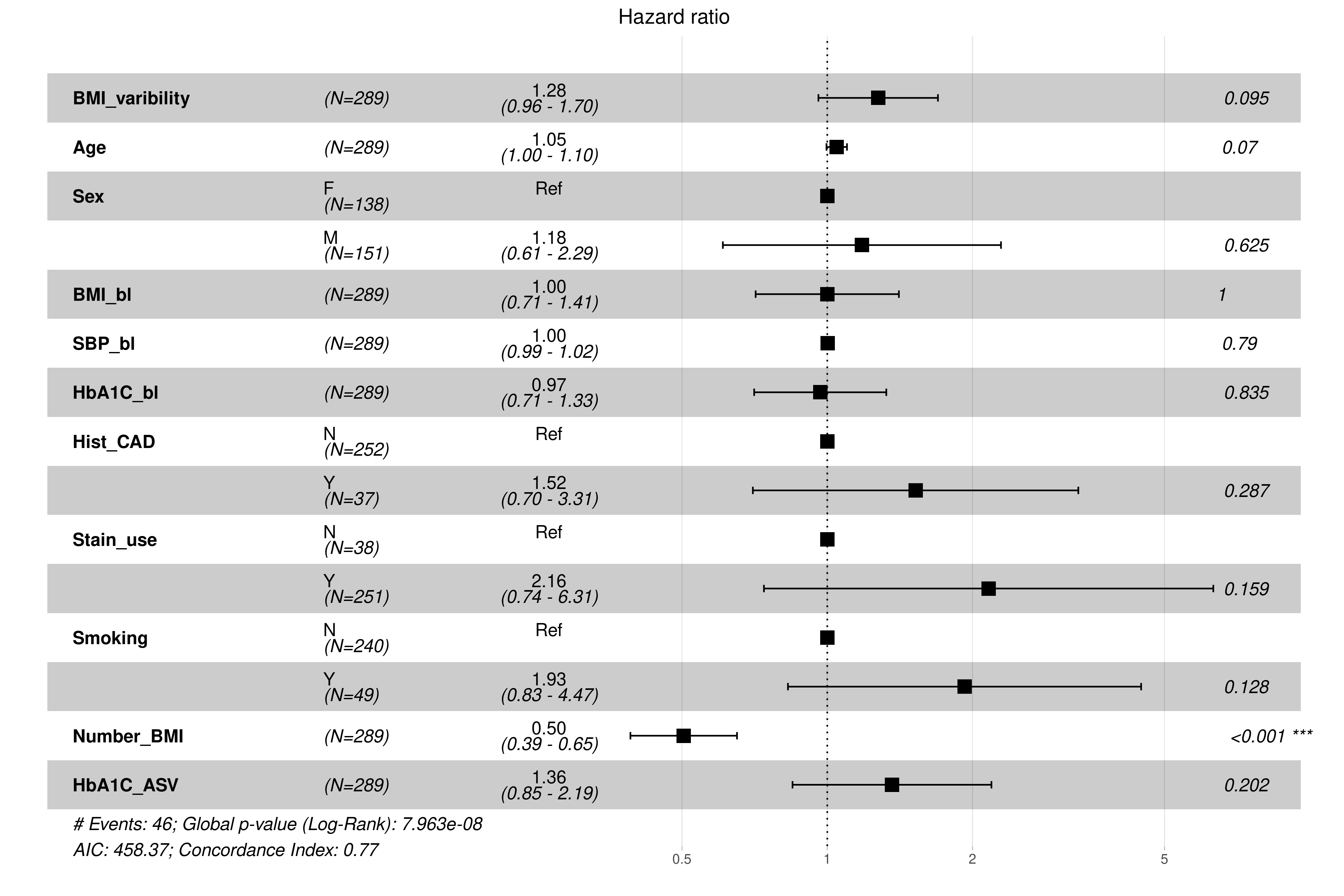

Figure S4(b)**:** *A forest plot summarising the adjusted hazard ratio (HR) and 95% confidence interval (CI) of 3-point major adverse cardiovascular events (MACE) risk associated with a +1 standard deviation (SD) increase in BMI variability within the Tayside Bioresource (n = 6980) cohort in individuals with normal baseline BMI after adjustment for age, sex, baseline BMI, systolic blood pressure, baseline HbA1c, history of coronary artery disease, statin use, smoking, number of BMI measurements, and ASV HbA1c. Risk estimates for each covariate are calculated via multivariate Cox regression.*

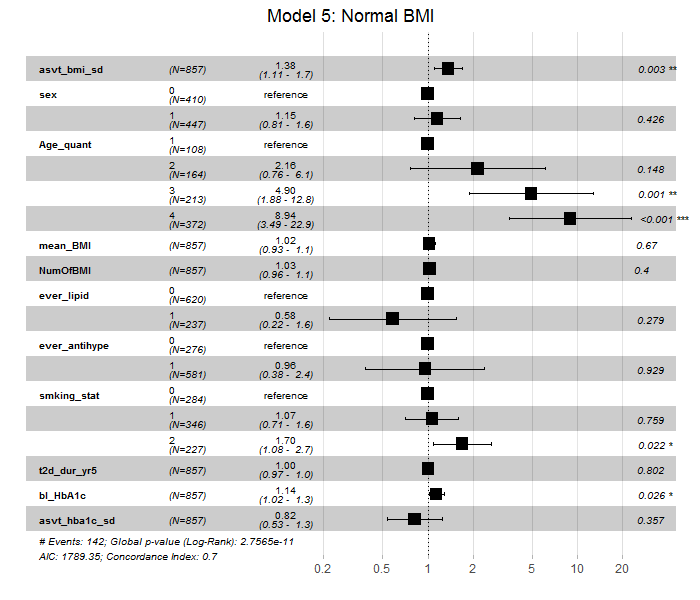

Figure S5(a)**:** *A forest plot summarising the adjusted hazard ratio (HR) and 95% confidence interval (CI) of 3-point major adverse cardiovascular events (MACE) risk associated with a +1 standard deviation (SD) increase in BMI variability within the REWIND (n = 4440) trial placebo-arm cohort in individuals with overweight baseline BMI after adjustment for treatment, baseline BMI, sex, age, smoking, type 2 diabetes duration, systolic blood pressure, statin use, baseline HbA1c, number of BMI measurement, and ASV HbA1c. Risk estimates for each covariate are calculated via multivariate Cox regression.*

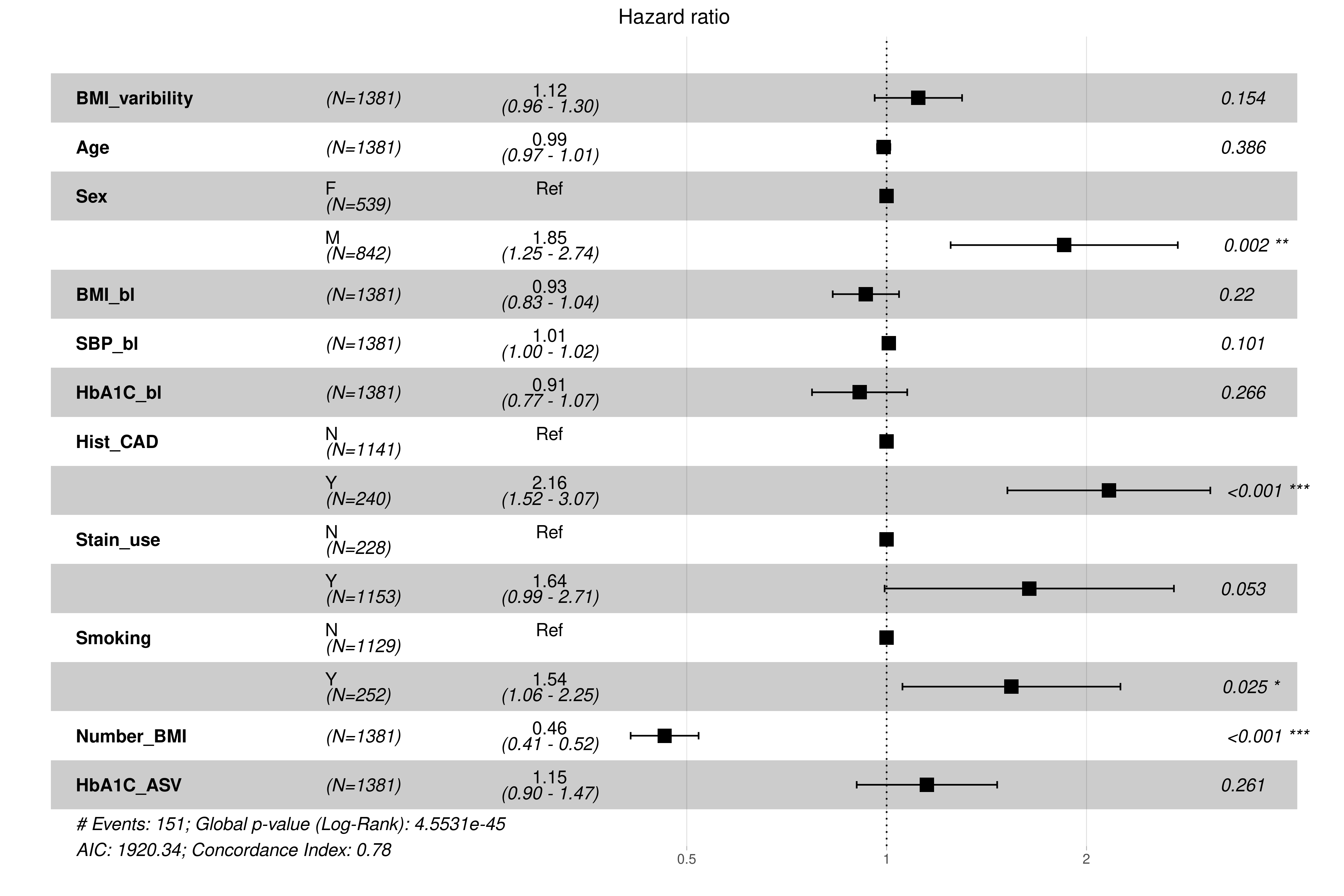

Figure S5(b)**:** *A forest plot summarising the adjusted hazard ratio (HR) and 95% confidence interval (CI) of 3-point major adverse cardiovascular events (MACE) risk associated with a +1 standard deviation (SD) increase in BMI variability within the Tayside Bioresource (n = 6980) cohort in individuals with overweight baseline BMI after adjustment for age, sex, baseline BMI, systolic blood pressure, baseline HbA1c, history of coronary artery disease, statin use, smoking, number of BMI measurements, and ASV HbA1c. Risk estimates for each covariate are calculated via multivariate Cox regression.*

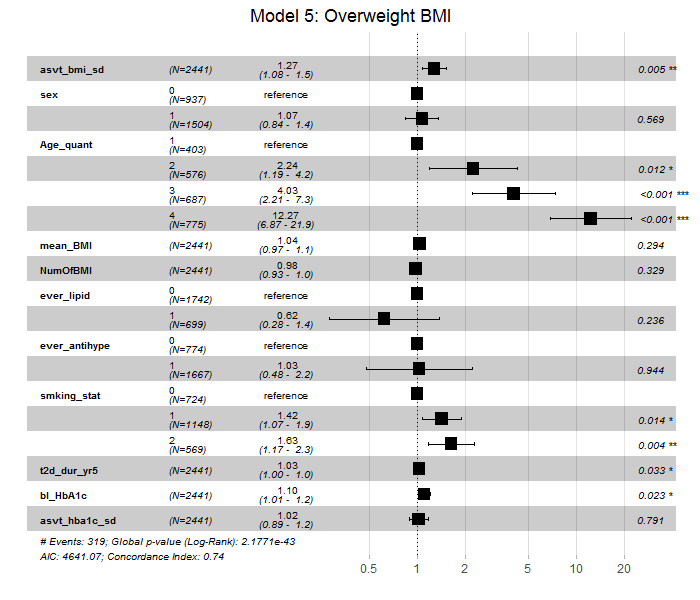

Figure S6(a)**:** *A forest plot summarising the adjusted hazard ratio (HR) and 95% confidence interval (CI) of 3-point major adverse cardiovascular events (MACE) risk associated with a +1 standard deviation (SD) increase in BMI variability within the REWIND (n = 4440) trial placebo-arm cohort in individuals with obese baseline BMI after adjustment for treatment, baseline BMI, sex, age, smoking, type 2 diabetes duration, systolic blood pressure, statin use, baseline HbA1c, number of BMI measurement, and ASV HbA1c. Risk estimates for each covariate are calculated via multivariate Cox regression.*

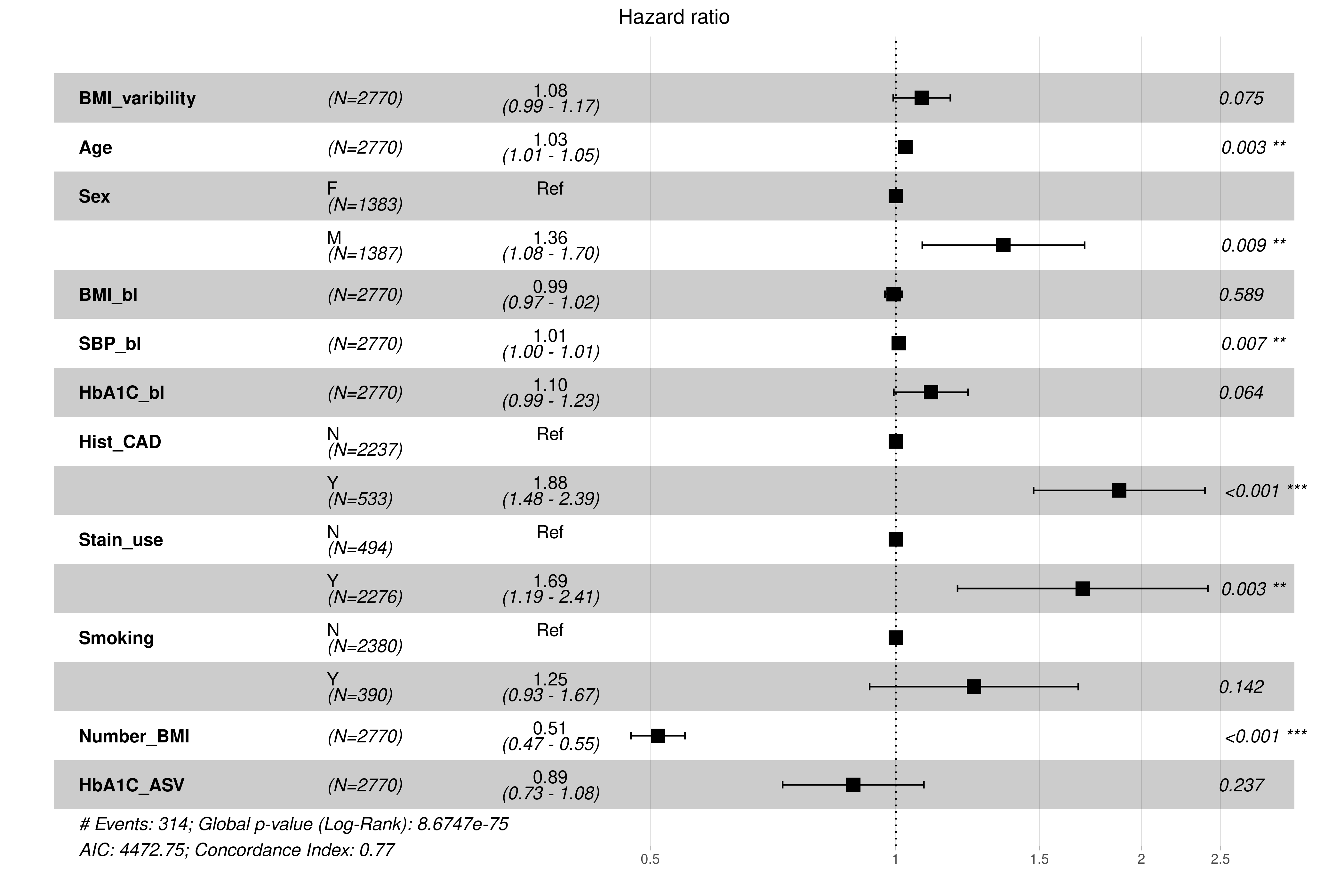

Figure S6(b)**:** *A forest plot summarising the adjusted hazard ratio (HR) and 95% confidence interval (CI) of 3-point major adverse cardiovascular events (MACE) risk associated with a +1 standard deviation (SD) increase in BMI variability within the Tayside Bioresource (n = 6980) cohort in individuals with obese baseline BMI after adjustment for age, sex, baseline BMI, systolic blood pressure, baseline HbA1c, history of coronary artery disease, statin use, smoking, number of BMI measurements, and ASV HbA1c. Risk estimates for each covariate are calculated via multivariate Cox regression.*

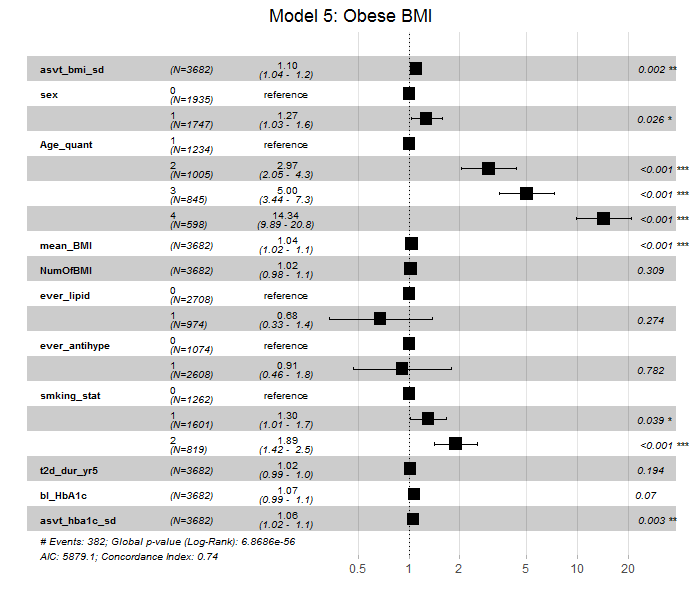

### Meta-analysis results

Figure S7**:** *A forest plot showing the summative risk of 3-point major adverse cardiovascular events (MACE) risk for individuals within quartile 4 (using quartile 1 as reference) of BMI variability.* *The results of the analyses of the Harmony Outcomes (n = 9198), REWIND (n = 4440), and EMPA-REG OUTCOME (n = 2333) are meta-analysed here using a fixed effect model.*

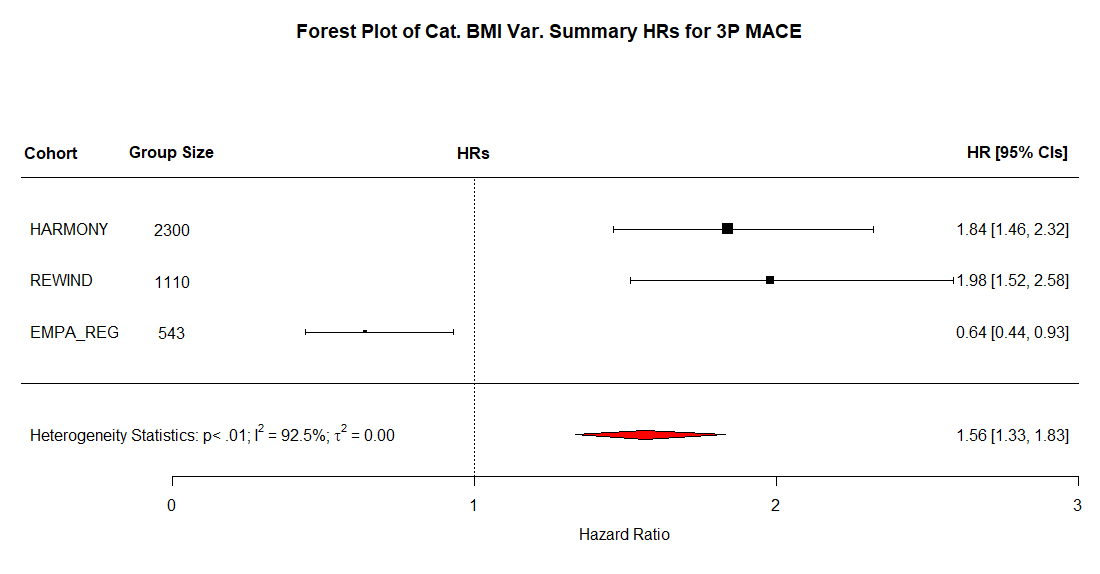

Figure S8: *A forest plot showing the summative risk of 3-point major adverse cardiovascular events (3P-MACE) risk associated with a +1 standard deviation (SD) increase in BMI variability within the clinical trial cohorts. The results of the analyses of the* Harmony Outcomes (n = 9198), REWIND (n = 4440), and EMPA-REG OUTCOME (n = 2333) *are meta-analysed here using a fixed effect model.*

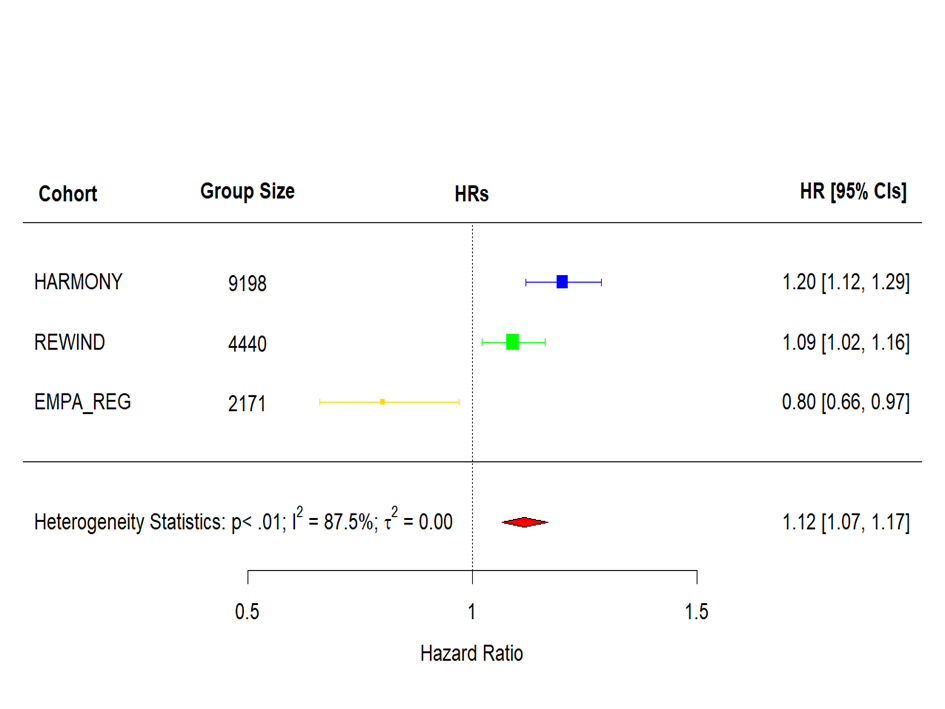

Figure S9: *A forest plot showing the summative risk of 3-point major adverse cardiovascular events (3P-MACE) risk associated with a +1 standard deviation (SD) increase in BMI variability within all cohorts. The results of the analyses of the Harmony Outcomes (n = 9198), REWIND (n = 4440), EMPA-REG OUTCOME (n = 2333), and Tayside Bioresource (n = 6980)* *are meta-analysed here using a fixed effect model.*

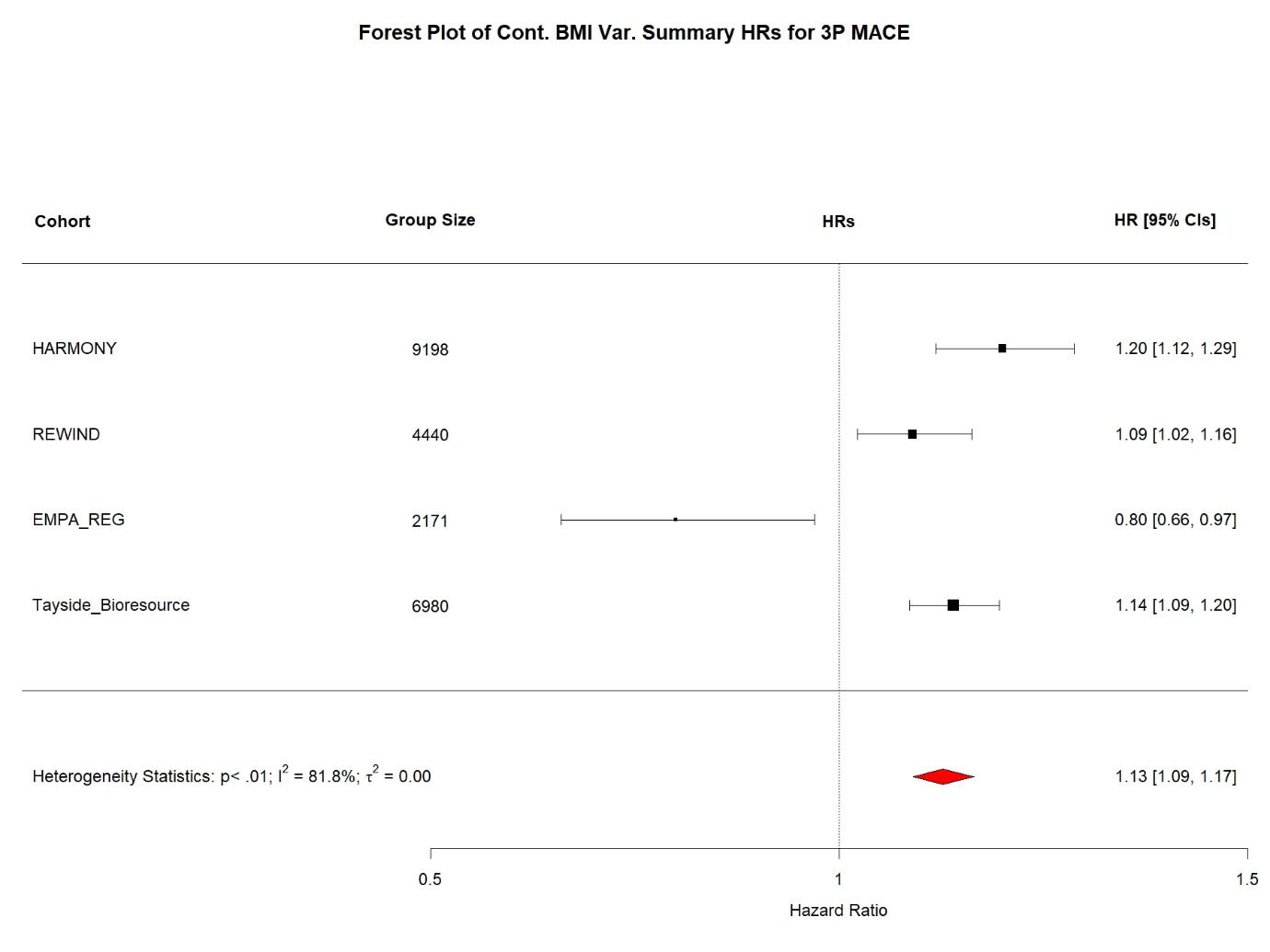

Figure S10: *A forest plot showing the summative risk of 3-point major adverse cardiovascular events (MACE) risk for individuals within quartile 4 (using quartile 1 as reference) of BMI variability.* *The results of the analyses of the Harmony Outcomes (n = 9198), REWIND (n = 4440), EMPA-REG OUTCOME (n = 2333), and Tayside Bioresource (n = 6980) are meta-analysed here using a fixed effect model.*

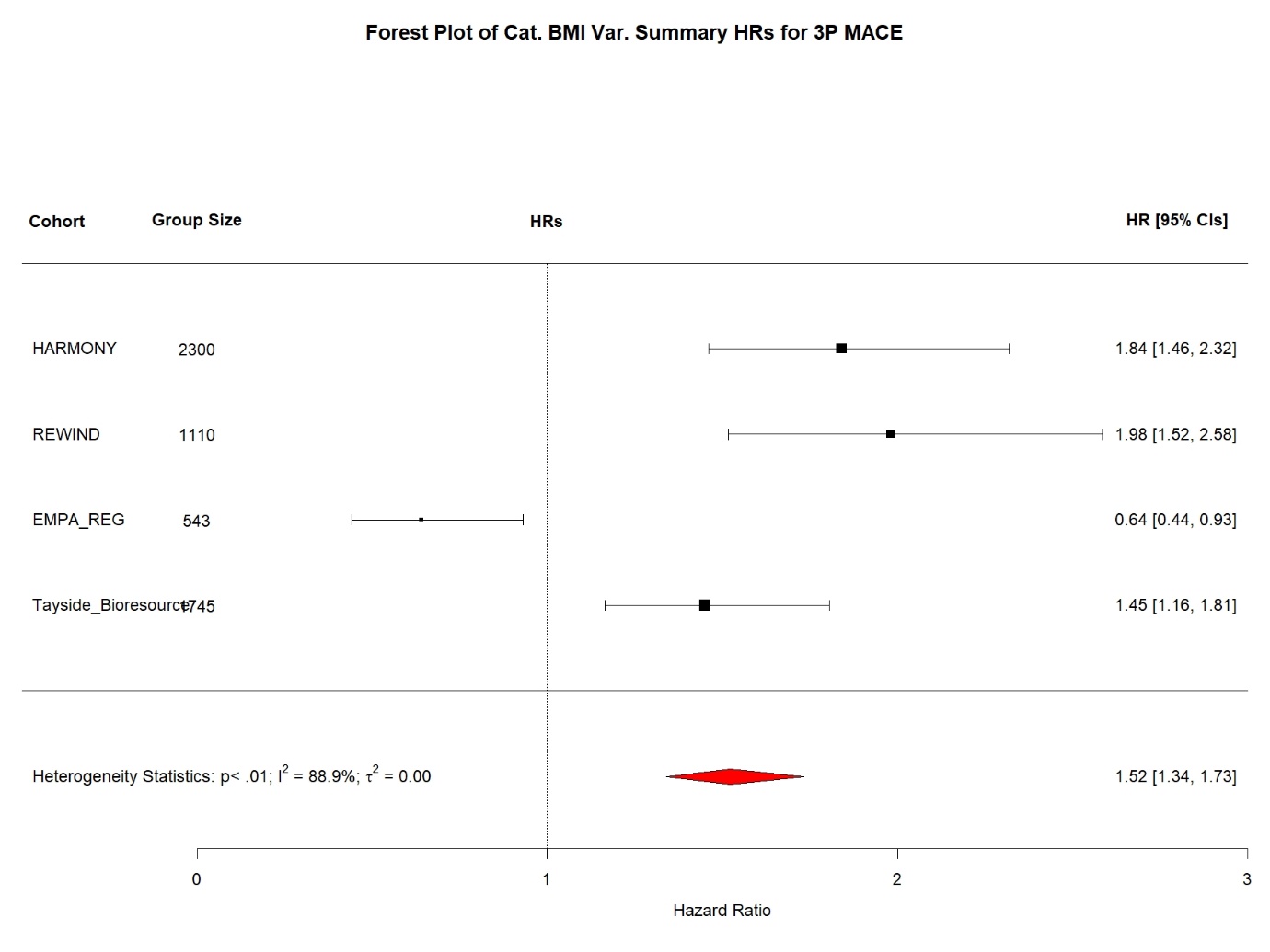

### Example of BMI trends of patients with high and low variability

Figure S11**:** *Spaghetti plots of BMI trendlines from 10 randomly selected individuals from the Tayside Bioresource (n = 6980) cohort.* Trendlines in pink represent the BMI trendlines of individuals from the most variable quartile (quartile 4). Trendlines in green represent the BMI trendlines of individuals from the least variable quartile (quartile 1).

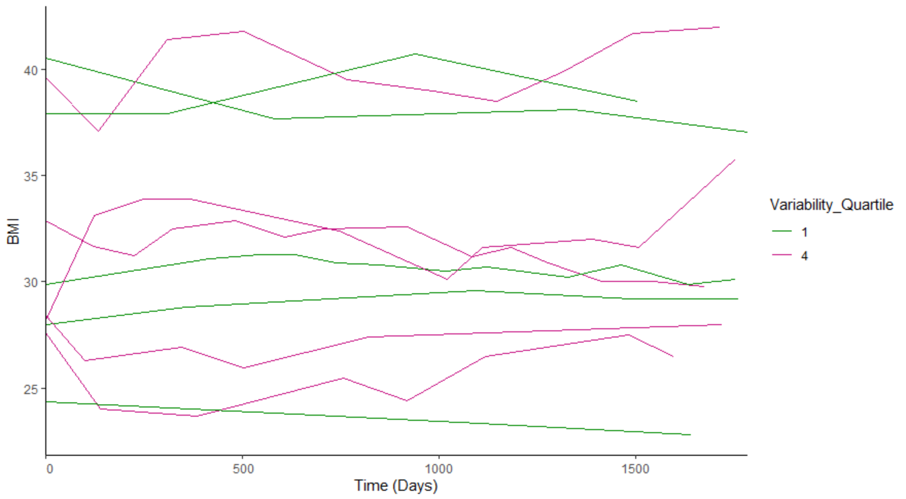

### References

1.	Green JB, Hernandez AF, D'Agostino RB, Granger CB, Janmohamed S, Jones NP, et al. Harmony Outcomes: A randomized, double-blind, placebo-controlled trial of the effect of albiglutide on major cardiovascular events in patients with type 2 diabetes mellitus-Rationale, design, and baseline characteristics. Am Heart J. 2018;203:30-8.

2.	Gerstein HC, Colhoun HM, Dagenais GR, Diaz R, Lakshmanan M, Pais P, et al. Design and baseline characteristics of participants in the Researching cardiovascular Events with a Weekly INcretin in Diabetes (REWIND) trial on the cardiovascular effects of dulaglutide. Diabetes Obes Metab. 2018;20(1):42-9.

3.	Zinman B, Inzucchi SE, Lachin JM, Wanner C, Ferrari R, Fitchett D, et al. Rationale, design, and baseline characteristics of a randomized, placebo-controlled cardiovascular outcome trial of empagliflozin (EMPA-REG OUTCOME™). Cardiovascular Diabetology. 2014;13(1):102.

4.	McKinstry B, Sullivan FM, Vasishta S, Armstrong R, Hanley J, Haughney J, et al. Cohort profile: the Scottish Research register SHARE. A register of people interested in research participation linked to NHS data sets. BMJ Open. 2017;7(2):e013351.

5.	Hébert HL, Shepherd B, Milburn K, Veluchamy A, Meng W, Carr F, et al. Cohort Profile: Genetics of Diabetes Audit and Research in Tayside Scotland (GoDARTS). International Journal of Epidemiology. 2017;47(2):380-1j.
